# The spatiotemporal distribution of substandard and falsified antimalarial medicines in Africa, 1996-2019

**DOI:** 10.64898/2025.12.22.25342680

**Authors:** Lawrence B. Adipo, Jessica A. Mendes, Ngan Thi Do, Kerlijn Van Assche, Paula Moraga, Stella Maris Nanyonga, Nicole Stoesser, Christiane Dolecek, Paul N. Newton, Ben S. Cooper, Celine Caillet, Sean M. Cavany

## Abstract

In 2024, an estimated 282 million malaria cases and 610,000 deaths occurred globally, many attributable to inadequate access to quality-assured and efficacious antimalarials. Africa bears the greatest burden of malaria, yet the spatial and temporal distribution of substandard and falsified (SF) antimalarials across the continent remains poorly understood. We extracted data from the Infectious Disease Data Observatory (IDDO) Medicine Quality Scientific Literature Surveyor database on SF antimalarial prevalence. We applied spatiotemporal modelling to estimate the proportion of antimalarials that were SF by country and year. We constructed three different models with identical spatial structures and covariates but different specifications of the temporal trends. We modelled spatial effects using the Besag-York-Mollié model (BYM). We fitted the models using integrated nested Laplace approximation (INLA) and compared models’ predictive ability using the widely applicable information criterion (WAIC). We extracted data from 76 random and convenience surveys conducted between 1996 and 2019, including 8,213 antimalarial samples in total. The model with the best predictive performance included an interaction between space and time (WAIC=499.430), suggesting that the highest SF prevalence occurred between 1996 and 2003, with higher predictions in West and Central Africa. Most countries had no clear temporal patterns, but a notable decline was estimated in Kenya, Uganda, Tanzania, and Madagascar beginning around 2003. This study provides country-level, time-varying estimates of SF antimalarial prevalence, improving understanding of spatial and temporal heterogeneity in Africa. Although these findings can inform targeted interventions, considerable uncertainty reflects sparse surveillance data and warrants cautious interpretation.

## INTRODUCTION

Globally, it is estimated that there were almost 282 million malaria cases and 610,000 malaria deaths in 2024. The number of estimated malaria cases and deaths increased by 9 million (3.5%) and 12,000, respectively, compared with 2023.^1^ The WHO African region bears much of the burden of malaria; in 2024, it accounted for 88% of global malaria cases and 95% of malaria deaths. Many of these deaths would be avoidable if patients had ready access to quality-assured and efficacious antimalarials. ^2^

Similarly, the African continent also has a high burden of substandard and falsified (SF) medicines, according to scientific studies and regular WHO SF medical product alerts.^3–5^ The World Health Organization (WHO) defines substandard medicines as authorized medical products that fail to meet either quality standards, specifications, or both.^6^ They can result from poor manufacturing practices, inadequate quality assurance, shipping, storage conditions, or sale of the medicine beyond the expiration date.^7^ Falsified medicines are defined as medical products that deliberately or fraudulently mispresent their identity, composition, or source.^6^ Poor-quality antimalarials substantially impact public health, contributing to treatment failures that extend illness duration, prolong hospital stays, contribute to antimalarial resistance, and likely increase malaria mortality.^6–8^ These health impacts in turn have important economic consequences. For instance, they impose considerable financial burdens on patients, their families, and healthcare systems due to increased treatment costs and decreased productivity, ^7, 9^ while also exacerbating health inequalities.^10^

Despite these impacts, our understanding of SF antimalarial prevalence patterns through time and space remains very limited. The global prevalence of SF antimalarials has been estimated at 19.1% (95% CI, 15.0%-23.3%).^11^ Estimates of SF medical product prevalence are based on studies that detect SF medicines using a range of strategies, including visual inspection of packaging and more complex technologies. However, the prevalence of SF antimalarials is not uniform, but instead shows substantial variation across different locations and over time. This spatiotemporal heterogeneity is often obscured when estimates are aggregated into regional or global prevalence figures, masking local patterns and undermining efforts to target interventions effectively.

In this study, we aim to address some of these gaps using a spatiotemporal modelling approach. We synthesize data from published surveys of antimalarial quality in African countries where at least one survey was conducted in the period 1996 - 2019. We additionally leverage data on a range of covariates plausibly associated with the prevalence of SF antimalarials and estimate spatial random effects and temporal patterns. This allows us to produce estimates of the prevalence of SF antimalarials in each year from 1996 to 2019 for all included countries. These spatiotemporal estimates offer new insights into the prevalence of SF antimalarials and provide evidence to guide targeted interventions, including strengthening medicine regulatory systems, promoting regional regulatory collaboration, and enhancing post-marketing surveillance, thereby contributing to global access to good-quality medicines. ^12^

## MATERIALS AND METHODS

### Data Sources

#### i) SF antimalarials

We retrieved data from the ‘Medicine Quality Scientific Literature Surveyor’ (hereafter ‘the Surveyor’) database that was developed by the Medicine Quality Research Group (MQRG) and the Infectious Diseases Data Observatory (IDDO) at the University of Oxford.^13^ The Surveyor was built as an extension to the original ‘WorldWide Antimalarial Resistance Network Antimalarial Quality Surveyor’.^14^ The Surveyor’s data originate from systematic reviews of the scientific literature on the quality of medical products, and the database is regularly updated with new scientific studies on those SF medical products identified by the MQRG via PubMed and Google Scholar.^3, 14–19^ All samples collected from random (i.e. samples were collected from outlets selected following randomization procedures) and convenience sampling approaches (i.e. samples were collected from outlets selected with a non-probability sampling method) were included.^20^ We extracted the name of the country and the year in which the survey was conducted and published, the name of the stated active pharmaceutical ingredient (API), the type of outlet where the samples were collected, the sampling approach (convenience vs random sampling of the outlets), the number of samples collected and the number of samples that were found to be SF (i.e. failing at least one of the quality tests that was conducted). We used the definitions for substandard and falsified antimalarials as used in each individual paper.

#### ii) Covariate data

We obtained data on several other variables that could plausibly help predict the prevalence of SF medicines, including World Bank governance indicators (government effectiveness, rule of law, regulatory quality, control of corruption, voice and accountability, and political stability),^21^ malaria incidence per 1000 person,^22^ GDP per capita,^23^ and total population^23^ (S1&2: Online supplementary, Table 1&2). We extracted data on each of these covariates for each country and year under study.

### Data processing

Data were imported into R software version 4.4.0.^24^ Five studies conducted before 1996 were excluded as there were no covariate data from these years. We also defined a neighbourhood matrix (as explained under the data analysis section) for countries, indicating which countries share a border. We excluded surveys when only one sample in the country was tested and results for countries whose bordering countries had no samples tested for quality, as there is insufficient information for the Besag, Yorke, and Mollie (BYM) model to estimate prevalence for those countries. This resulted in the exclusion of one sample from Comoros and one from Morocco.

### Data analysis

#### i. Descriptive statistical analysis

Categorical variables are expressed as counts and percentages, and we present summary statistics within strata, in maps for each year under study, and as time series plots for each country.

#### ii. Spatiotemporal modelling

Before fitting the models, we performed multicollinearity analysis of the predictor variables using the variance inflation factor (VIF). The *vif* function in the *car* package version 3.1.2 in R software was used to generate the VIF values.^25^ Predictors with VIF greater than 10 were considered to have a high degree of multicollinearity and dropped from the final model.^26^Also, correlation analysis was conducted to assess the association between the outcome variable and predictors, using the *corrplot* ^27^ package version 0.95 to construct the correlation matrix (S5: Online supplementary, Figure 2).

To obtain estimates of SF proportions for each country and year of study, we fitted three Bayesian spatiotemporal models. These models used a logit link function and assumed a binomial likelihood for the number of antimalarials that were SF in a particular country and year,

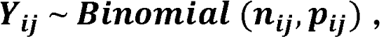

where *Y_ij_* is the observed number of SF samples in country *i* and year *j*, *n*_iJ_ is the total number of samples, and *p_ij_* is the estimated proportion of samples that are SF. In addition to the covariates mentioned above, we included spatial effects according to the BYM model.^28^ The BYM model includes two spatial random effects: one which incorporates spatial structure using an intrinsic conditional autoregressive function (iCAR; see below for equation); and one that is unstructured.

Specifically, the Bayesian spatiotemporal model assumes that the underlying prevalence of SF antimalarials varies smoothly over space and time through structured spatial and temporal random effects. This allows neighbouring countries and adjacent years to contribute partial information to one another, while not assuming that they share identical prevalence. Where observed data support differences between neighbouring countries or years, the model allows these differences to remain. Furthermore, the model naturally reflects greater uncertainty in settings where survey data are sparse.

The first model included a parametric country-level time trend based on the Bernardinelli model.^29^ The parametric formulation assumes that the linear predictor can be written as

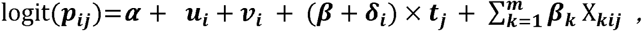

where *α* represents the overall SF proportion in Africa, *Х_kíj_* represents the *k* predictor variables, *v_í_* is a country-level spatial random effect modelled using an iCAR distribution, *r*_i_ is an unstructured exchangeable component that models uncorrelated noise, *v_i_ ~ N(0, σ^2^_u_)*, *β* is a global linear trend effect, and *s*_i_ is a fixed interaction between space and time representing the difference between the global trend *β* and the area-specific trend. The iCAR distribution assigned to the spatial random effect mentioned above shares information between neighbours, 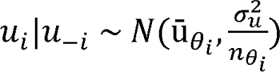 where *θ_i_,n_θi_* represents the set of neighbours and number of neighbours of country *i* respectively. In this study, we define countries to be neighbours if they share a common boundary. The neighbourhood matrix was constructed using *poly2nb* function of the *spdep R* package version 1.3.3. ^30^

The second model included a non-linear random walk for the temporal trend in each country. This is a nonparametric formulation of the linear predictor,^31^ written as

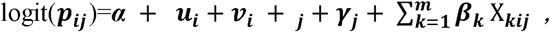

Here *α*, *u*_i_, and *r*_i_ have the same parameterization as in the first model equation above, *y*_J_ represents the temporally structured effect, modelled using a Random Walk of Order 1 (RW1) and defined as

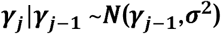

Finally, the term *Φ_j_* is specified using Gaussian exchangeable prior: *Φ_j_ ~ N(0, 1/τ_Φ_*.

The third model was an extension of the second model but including an interaction term between space and time *(δ_ij_), δ_ij_ ~ N(0, 1/τ_Φ_*. The interaction term explains the differences in the time trend of the SF proportions for different countries. The formulation of this mode is specified as

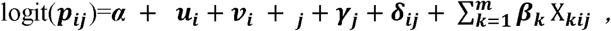

We fitted our models to the prevalence of SF antimalarials using the integrated nested Laplace approximation (INLA) method for approximate Bayesian inference, as implemented in the *R-INLA* package version 24.5.10.^32^

We compared models using the Deviance Information Criteria (DIC) ^33^ and Watanabe Akaike Information Criteria (WAIC).^34, 35^ Both metrics include a measure of goodness-of-fit and a penalization for complexity. Models with lower DIC and WAIC are better supported by the data, in the sense that they are expected to have better out-of-sample predictive accuracy.

We present our results using maps of the posterior mean and 95% credible intervals (CrI), and time series plots for each country. All plots and maps were created with the *ggplot2* package version 3.5.1.^36^

#### iii. Sensitivity analysis

To assess the influence of very small surveys on the model estimates, we conducted a sensitivity analysis excluding surveys with small sample sizes (fewer than 11 tested medicine samples). Model performance was compared with the primary analysis using the Deviance Information Criterion (DIC) and the Widely Applicable Information Criterion (WAIC). All the datasets and code used are available at https://github.com/babulawrence/SF-Antimalarials-in-Africa.

## RESULTS

### Baseline characteristics of the antimalarial samples

In total, 76 surveys on quality of antimalarials conducted between 1996 and 2019 were included (Figure 1). Of these, 52 (68.4%) used a convenience sampling approach and 24 (31.6%) used random sampling. A total of 8213 samples were included, of which 1782 (21.7%) were SF. More than half of all samples (4972 / 82,13; 60.5%) and more than half of SF samples (1371/1782; 76.9%) were from convenience surveys. The proportion of samples which were SF was higher in convenience surveys (1371/4972; 27.6%) than in random surveys (411 / 3241; 12.7%).

**Figure 1:**
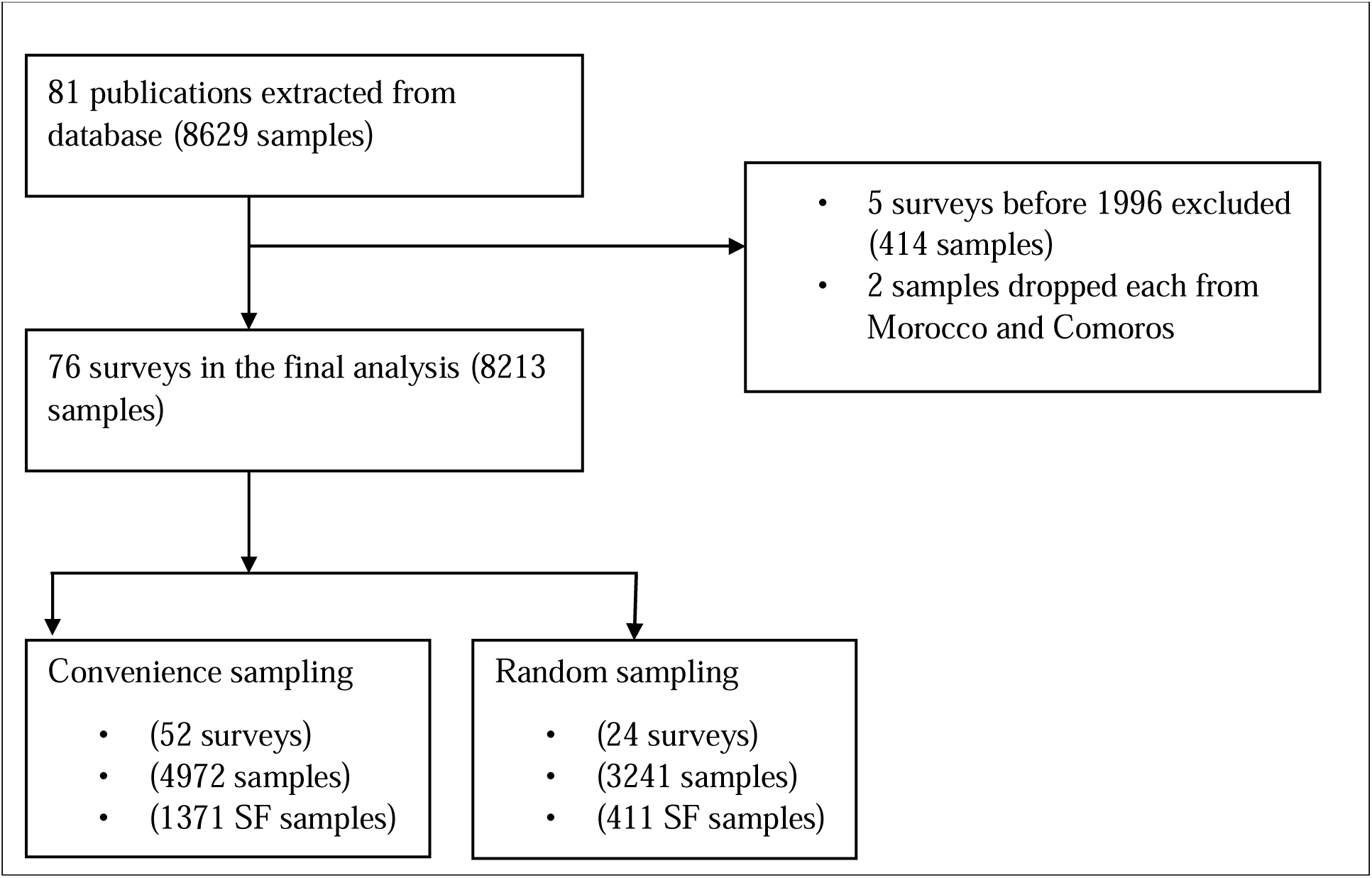
Data processing flowchart

Most samples (5207 / 8213; 63.4%) came from an unspecified combination of outlets (i.e. the samples were collected from different types of outlet but results were not broken down by type of outlet), followed by private pharmacies (1488 / 8213; 18.1%) (Table 1). The highest number of SF samples (924 / 1782; 51.9%) was reported in unspecified combinations of outlets followed by private pharmacies (358 / 1782; 20.1%). The highest proportion of SF antimalarials was identified in ‘other outlets’ (71.4%) followed by wholesalers (46.6%) and unknown outlets (43.2%). Health centres (19.2%) and combinations of outlets (17.7%) exhibited the lowest proportions.

**Table 1:**
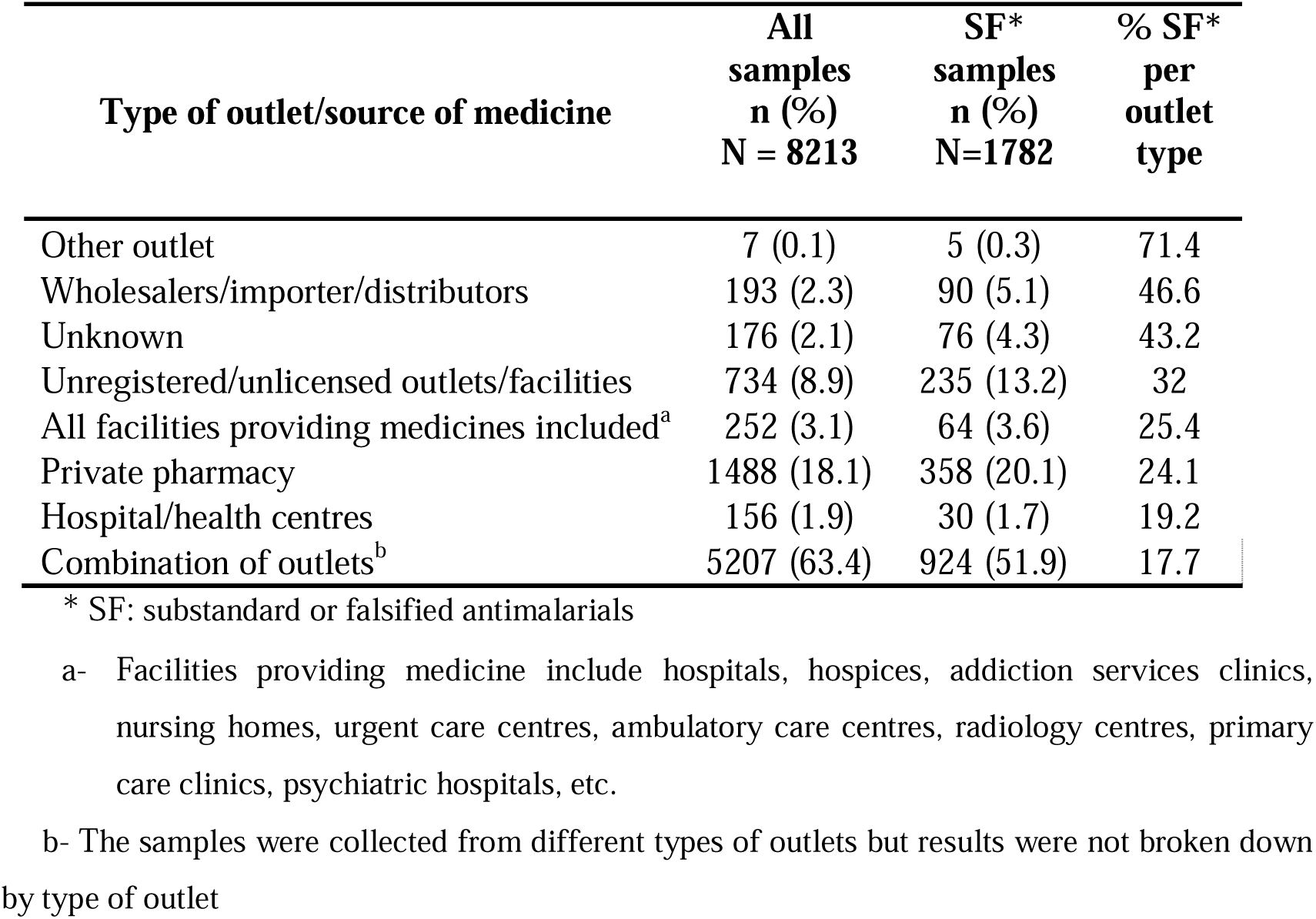
Numbers and proportions of samples collected and SF by outlet types.

Of the 8213 antimalarials, most were stated to be combinations of artemether and lumefantrine (2,063; 25.1%), followed by quinine (1595; 19.4%) (Table 2). The highest percentages of SF among medicines with ten or more samples were found for samples stated to be artesunate and sulphadoxine-pyrimethamine combinations (87.0%), tetracycline (66.7%) and artemether (59.1%).

**Table 2:** Classification of antimalarial drugs.

| Stated *API/API combination | All samples<br>n (%)<br>N=8213 | SF samples<br>n (%)<br>N=1782 | % SF°<br>per<br>API |
| --- | --- | --- | --- |
| Halofantrine | 1 (0) | 1 (0.1) | 100.0 |
| Artesunate+Sulphadoxine-Pyrimethamine | 100 (1.2) | 87 (4.9) | 87.0 |
| Tetracycline | 15 (0.2) | 10 (0.6) | 66.7 |
| Artemether | 22 (0.3) | 13 (0.7) | 59.1 |
| Artesunate-Amodiaquine Co-formulated | 228 (2.8) | 112 (6.3) | 49.1 |
| Arteether | 17 (0.2) | 8 (0.4) | 47.1 |
| Dihydroartemisinin | 37 (0.5) | 17 (1.0) | 45.9 |
| Chloroquine | 669 (8.1) | 285 (16.0) | 42.6 |
| Artesunate-Amodiaquine Unknown formulation | 147 (1.8) | 59 (3.3) | 40.1 |
| Sulfamethoxazole-Pyrimethamine | 283 (3.4) | 112 (6.3) | 39.6 |
| Amodiaquine | 158 (1.9) | 61 (3.4) | 38.6 |
| Artesunate+Amodiaquine Co-blistered | 280 (3.4) | 108 (6.1) | 38.6 |
| Artesunate | 445 (5.4) | 132 (7.4) | 29.7 |
| Dihydroartemisinin-Piperaquine | 355 (4.3) | 95 (5.3) | 26.8 |
| Sulphadoxine-Pyrimethamine | 1045 (12.7) | 246 (13.8) | 23.5 |
| Artemether-Lumefantrine | 2063 (25.1) | 246 (13.8) | 11.9 |
| Artemisinin-unspecified partner medicine | 309 (3.8) | 31 (1.7) | 10.0 |
| Quinine | 1595 (19.4) | 144 (1.8) | 9.0 |
| Dihydroartemisinin-Sulfadoxine-Pyrimethamine | 444 (5.4) | 15 (0.8) | 3.4 |

\* API: Active pharmaceutical ingredient; °SF: Substandard or falsified antimalarials

### Proportion of SF antimalarials by country

#### i) Observed data

Samples collected from 27 countries in Africa were included in this study. In most countries, either one (8/27; 29.6%), two (6/27; 22.2%), or 3 (6/27; 22.2%) surveys only were identified during the 24 years. The largest number of surveys was in Nigeria (n=13). Overall, 2008 was the year with the highest number of surveys identified (n=16), and we observed a decline in the number of surveys per year after 2009, except for a peak in 2014 (n=8). No country had surveys for each of the 24 consecutive years under study (S3: Online supplemental, Figure 1). Over the years, SF proportions were reported intermittently with peaks around 1997, 2004-2006, 2008, 2012, and 2019. West and Central Africa accounted for most high-proportion years (Figure 2).

**Figure 2:**
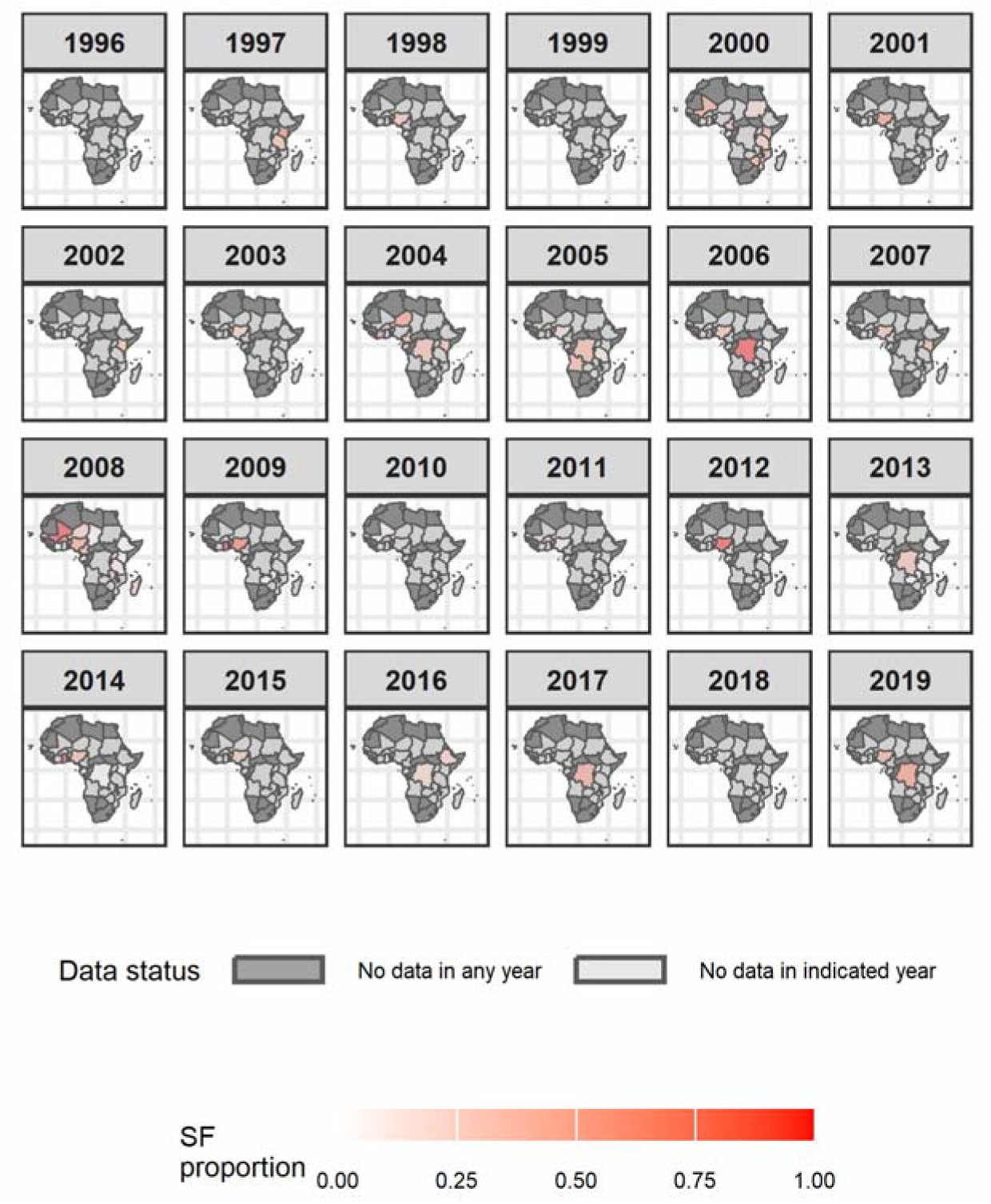
Proportion of antimalarials that were SF per year. For countries in dark grey there were no surveys in any year, while light grey represents countries with data for at least one year during the study period. The figure illustrates the spatial and temporal distribution of the surveys included in the analysis, highlighting substantial gaps in medicine quality surveillance across sub-Saharan Africa. This provides important context for interpreting the modelled prevalence estimates presented in subsequent figures. Detailed survey characteristics and sample counts for each country and year are provided in S8: Online supplementary, Table 4. Alt text: Maps showing observed SF proportions of antimalarials in Africa, 1996-2019

#### ii) Spatiotemporal modelling

Spearman correlation analysis showed that the World Bank governance indicators strongly correlated with each other (r>0.60). National GDP per capita had negligible correlation with both malaria incidence (r=-0.08) and population (r=0.06). Correlation between malaria incidence and population was also negligible (r=0.03). We observed a negligible correlation between SF prevalence and all the predictors (r<0.3) except for malaria incidence which showed low positive correlation (r=0.43) (S5: Online supplementary, figure 2). As indicated by the results from the VIF analysis (S4: Online supplementary, Table 3), no variables showed extreme multicollinearity so we include all as covariates.

Model 3, which had a random walk and a spatiotemporal interaction term, had the lowest DIC (486.280) and WAIC (499.430) (Table 3). For this reason, we proceeded with this model for our baseline analysis. Results from models 1 and 2 are included in the supplementary material (S7: Online supplementary, figure 4, S7: Online supplementary, Figure 5). Model 3 gave higher predicted SF proportions between 1996 and 2003 compared to subsequent years (Figure 3, S6: Online supplementary, Figure 3). Throughout the years under study, the highest predictions were mainly observed in West Africa, especially Niger, Nigeria, Ghana, Mali, and Côte d’Ivoire, and in the Democratic Republic of the Congo.

**Figure 3:**
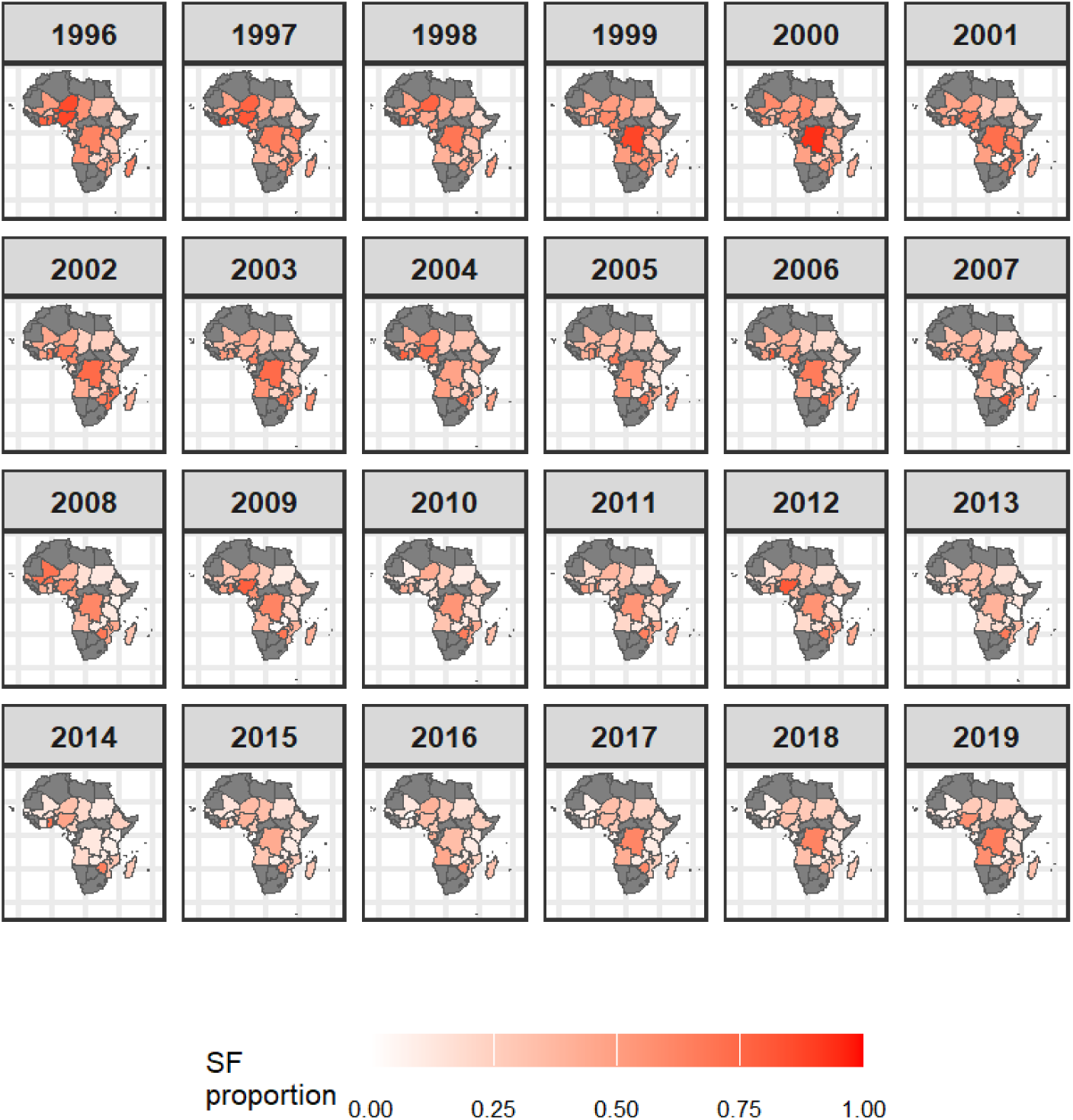
Maps showing posterior mean predicted proportion of antimalarials that are SF in Africa from 1996 to 2019. Maps showing 95% credible intervals of the predicted proportions in S6: Online supplementary, Figure 3. *Alt text: Maps showing predicted proportions of SF antimalarials in Africa from 1996 to 2019*.

**Table 3:** Posterior estimates (mean, standard deviation (SD), quantiles) DIC, WAIC, and maximum log-likelihood (MLL) for the substandard and falsified antimalarials in Africa model (1996-2019)

| Model | Parameter | Mean | SD | 2.50% | 50% | 97.50% |
| --- | --- | --- | --- | --- | --- | --- |
| Model 1: Parametric time trend | DIC =666.220; WAIC=754.500 ; MLL=-405.630 |  |  |  |  |  |
|  | Intercept | -0.674 | 1.053 | -2.766 | -0.670 | 1.395 |
|  | Government effectiveness | -0.714 | 0.562 | -1.797 | -0.721 | 0.408 |
|  | Regulatory authority | -1.294 | 0.450 | -2.187 | -1.29 | -0.422 |
|  | Rule of law | 0.138 | 0.579 | -1.029 | 0.148 | 1.246 |
|  | Control of corruption | 2.338 | 0.608 | 1.174 | 2.327 | 3.561 |
|  | Voice and accountability | -0.512 | 0.493 | -1.470 | -0.516 | 0.467 |
|  | Political stability | 0.547 | 0.280 | -0.010 | 0.550 | 1.089 |
|  | Malaria incidence (per 10 persons) | 0.439 | 0.139 | 0.164 | 0.439 | 0.709 |
| | GDP (\$10<sup>-3</sup> per capita) | -0.070 | 0.168 | -0.389 | -0.075 | 0.273 |
|  | Population (millions) | 0.020 | 0.024 | -0.022 | 0.017 | 0.073 |
| Model 2: Random walk | DIC =653.150 ; WAIC=746.250 ; MLL=-404.430 |  |  |  |  |  |
|  | Intercept | -2.762 | 0.893 | -4.523 | -2.761 | -1.006 |
|  | Government effectiveness | -2.814 | 0.703 | -4.202 | -2.811 | -1.441 |
|  | Regulatory authority | 0.727 | 0.551 | -0.355 | 0.727 | 1.808 |
|  | Rule of law | 1.492 | 0.638 | 0.255 | 1.487 | 2.757 |
|  | Control of corruption | 2.126 | 0.689 | 0.783 | 2.123 | 3.489 |
|  | Voice and accountability | -2.793 | 0.517 | -3.823 | -2.787 | -1.793 |
|  | Political stability | 1.612 | 0.342 | 0.963 | 1.605 | 2.305 |
|  | Malaria incidence (per 10 persons) | 0.621 | 0.167 | 0.291 | 0.621 | 0.946 |
| | GDP (\$10<sup>-3</sup> per capita) | -0.163 | 0.115 | -0.383 | -0.165 | 0.070 |
|  | Population (millions) | 0.034 | 0.010 | 0.015 | 0.034 | 0.055 |
| Model 3: Random walk + spatiotemporal interaction | DIC =486.280; WAIC=499.430; MLL=-332.270 |  |  |  |  |  |
|  | Intercept | -2.448 | 0.837 | -4.194 | -2.414 | -0.893 |
|  | Government effectiveness | 0.938 | 0.982 | -1.086 | 0.974 | 2.775 |
|  | Regulatory authority | -0.511 | 0.838 | -2.161 | -0.514 | 1.151 |
|  | Rule of law | -0.590 | 1.132 | -2.780 | -0.604 | 1.675 |
|  | Control of corruption | -0.077 | 0.959 | -1.852 | -0.117 | 1.915 |
|  | Voice and accountability | -1.336 | 0.746 | -2.835 | -1.325 | 0.088 |
|  | Political stability | 0.951 | 0.481 | 0.029 | 0.942 | 1.919 |
|  | Malaria incidence (per 10 persons) | 0.470 | 0.199 | 0.108 | 0.460 | 0.885 |
| | GDP (\$10<sup>-3</sup> per capita) | -0.303 | 0.113 | -0.532 | -0.301 | -0.086 |
|  | Population (millions) | 0.012 | 0.009 | -0.003 | 0.012 | 0.032 |

Examining the patterns through time reveals annual variations in proportions of SF antimalarials at the country level (Figure 3). The distribution varies across the countries, with Kenya, Uganda, Tanzania, and Madagascar having notable apparent declines in SF proportions beginning around 2003. Both Burkina Faso and Mali exhibited a sharp increase in the apparent proportion of SF antimalarials between 2004 and 2012. Predictions in Côte d’Ivoire showed a consistent decline, though with a few peaks in 2004, 2006, and 2016. We observed decreasing then increasing predicted SF proportions in some countries, such as Burundi and Equatorial Guinea. Their predicted proportions fell before 2016 and rose in subsequent years. Countries, such as Rwanda and Malawi, showed the opposite pattern: an increase in the predicted SF antimalarial proportion before 2012, followed by a sharp decline in later years. Our analysis reports variable patterns in SF prevalence for Nigeria, Ghana, and Togo, which have several low and high peaks across the study period. As expected, we observed higher uncertainties in countries with fewer samples, such as Chad, Zimbabwe, Madagascar, Angola, and Benin (S6: Online supplementary, Figure 3).

There are no substantial qualitative differences in the temporal patterns predicted by the two random-walk models (model 3: Figures 4 and model 2: S7, online supplementary, Figure 5). However, results from the parametric model (model 1) yielded qualitatively different predictions (S7: Online supplementary, Figure 4). With this model, we estimated a declining trend in SF proportions across the years for all the countries except for Nigeria, the Democratic Republic of Congo, Senegal, and Ethiopia (S7: Online supplementary, Figure 4). A sensitivity analysis excluding surveys with fewer than 11 tested medicine samples produced similar spatiotemporal prevalence estimates to the primary analysis while improving model fit (S11: Online supplementary, Table 6).

**Figure 4:**
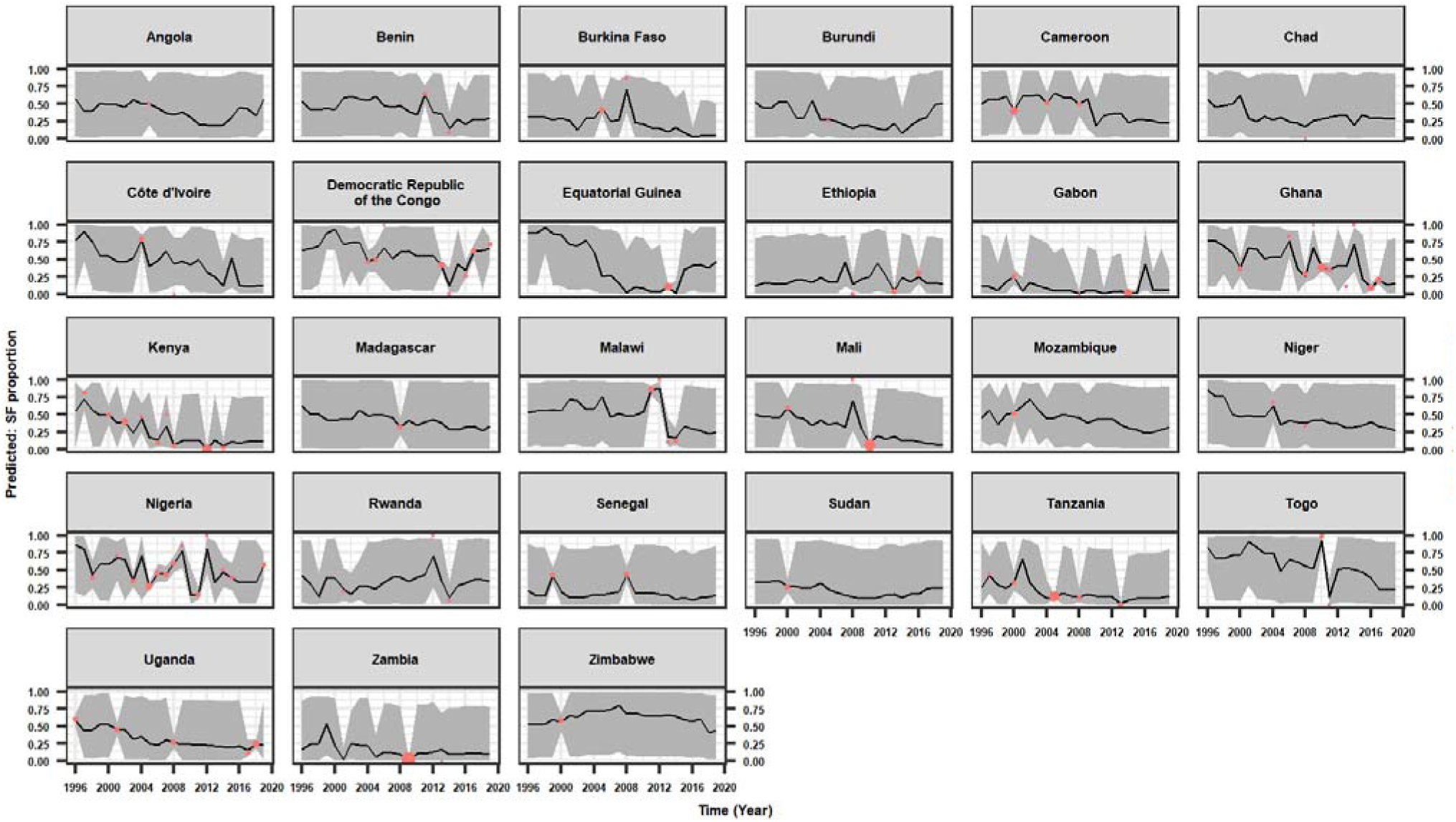
Posterior mean estimates (continuous dark line) from model 3 and their 95% credible intervals (grey area) showing trends in SF proportions in Africa. Data points in coral color represent the observed SF proportions. The size of the dots corresponds to the absolute sample size within each year. *Alt text: Plots of predicted mean estimates and their 95% credible intervals showing the trends in SF proportions of antimalarials in Africa from 1996 to 2019*

## DISCUSSION

Using a Bayesian spatiotemporal framework, we produced annual country-level estimates of the prevalence of SF antimalarials across Africa between 1996 and 2019 and provide new detail on their estimated spatiotemporal distribution. We estimated that SF antimalarials peaked during 1996–2003, with the highest prevalences in West and Central Africa. Most countries had no clear temporal patterns, but we found a notable decline in SF antimalarial prevalence in Kenya, Uganda, Tanzania, and Madagascar beginning around 2003. Conversely, Nigeria and the DRC experienced intermittent resurgences after 2009. The model with the best estimated predictive performance (as measured by WAIC) included a random walk temporal term and a space-time interaction, emphasizing the importance of allowing flexible prevalence trajectories that vary between countries.

Our overall estimate of SF prevalence for antimalarials of 21.7% (95% CI: 20.8%-22.6%) is consistent with general estimates for SF antimalarials from systematic reviews conducted by Ozawa et al (2018) that reported 19.7% (95% CI: 15.2% - 24.1%)^11^ and Mekonnen et al (2024) 15.6% (95% CI: 13.9%-16.2%).^37^ They also align with the pooled global prevalence estimate for SF antimalarials of 19.1% (95% CI: 15.0%-23.3%) by Ozawa et al (2022),^4^ but differ from the findings of Nayyar et al (2012), who reported a high estimated prevalence of 35% of SF in Africa.^2^ The determination of the accurate prevalence of SF medical products remains challenging due to limited surveys and different methodological approaches used in their estimation.^38^ Recent surveys have made greater use of risk-based post-market surveillance, which targets medicines, locations, or supply-chain settings considered at higher risk of quality problems. This approach is valuable for detecting SF products but may not provide estimates that are representative of the wider medicine market.^39^

Comparison of the three models shows that incorporating temporal and spatial interaction had a substantial impact on the estimated effects of the covariates. Although several governance indicators appeared strongly associated with the outcome in the simpler specifications, their magnitudes diminished and their uncertainty increased considerably in Model 3, which achieved the best predictive performance by a wide margin (lowest DIC and WAIC). This suggests that much of the apparent effect in the simpler models may have been due to unmodelled spatiotemporal patterns rather than representing genuine covariate effects, underscoring the importance of accounting for spatiotemporal interaction. Political stability is the only governance measure that remained positively associated with the outcome across all models and retained narrow intervals even under model 3. Malaria incidence also consistently showed a positive association, although its posterior uncertainty grew once spatiotemporal interaction was introduced. GDP per capita remained negatively associated across all models, with the most precise estimate under Model 3. In Model 3, the 95% credible intervals excluded zero for political stability, malaria incidence, and GDP per capita, whereas the intervals for the remaining governance indicators and population included zero (Table 3). However, the primary objective of this study was to develop a predictive spatiotemporal model and not to identify causal determinants of substandard and falsified antimalarials. Consequently, these regression coefficients should not be interpreted as evidence of causal relationships.

During the early included years, many African countries lacked sufficient national medicine regulatory frameworks to offer strict inspections and monitoring of medicines supplied within their countries and hence may have been unable to detect or prevent the rise of SF antimalarials.^2, 40^ Several African countries in the 1990s had poor governance, political instability, or economic instability, which led to collapse in the health systems and border insecurity. ^41–43^ These factors are potential drivers of the high prevalence of SF antimalarials in the 1990s. Subsequent declines in several countries may signal maturing regulatory frameworks and increased use of WHO pre-qualified suppliers. But the persistence of high-prevalence areas suggests that factors such as cross-border trade and insufficient regulatory capacity are still enabling the circulation of SF antimalarials.^2, 7, 38, 44–47^

Key strengths of our approach include our comprehensive inclusion of SF antimalarial surveys using the MQ surveyor database, our explicit modelling of spatial dependence using the BYM model, and our exploration of alternative temporal patterns. This study also has several important limitations. First, the spatial distribution of studies reporting on the quality of antimalarials in Africa was sparse. We analysed data from 27 countries, of which most countries reported only one or two surveys during the study period. The small number of surveys resulted in wide credible intervals in most countries. Also, we did not analyse SF antimalarials separately since many of the studies we included reported whether antimalarials were either substandard or falsified (SorF), and not which of these categories the medicine was within. These groups of medicines are likely to have different drivers, ^48^ and in future studies it would be important to look at the prevalence and drivers of these groups separately. We were only able to access publicly-accessible data; it is likely that national medicine regulatory authorities and the pharmaceutical industries have important datasets and better data sharing globally would help with better estimates and improve the data available to inform policy and interventions.^49^ With limited data, temporal patterns also often appear flat or with inconsistent patterns since it is difficult to detect gradual changes in SF proportions with insufficient data. Second, the predominance of convenience sampling (68.4% of surveys) may have led to bias in the estimates of SF prevalence compared with random designs. Third, unmeasured covariates could limit our predictive ability; for instance, governance indicators are unlikely to fully capture medicine quality regulatory performance. We did not analyze other potentially relevant factors, such as universal health coverage and health expenditure, because data for these variables were insufficient across the study period. Lastly, our country-level approach masks sub-national heterogeneity that is likely to be substantial, particularly in larger countries such as Nigeria and DRC. We were also unable to analyze differences between urban and rural regions because of the absence of location-specific information in these settings.

Although Bayesian hierarchical models provide a flexible framework for analysing sparse and heterogeneous datasets, the resulting estimates remain dependent on the quality and representativeness of the underlying medicine quality surveys. Consequently, the findings should be interpreted alongside the uncertainty intervals and in the context of the available evidence. Also, the model assumes that the included medicine quality surveys provide unbiased estimates of the underlying prevalence within their sampled populations. However, many surveys were conducted using convenience sampling strategies rather than nationally representative sampling. Although the Bayesian hierarchical framework accounts for sampling variability, it cannot fully correct for systematic bias in the underlying surveys. Consequently, prevalence estimates should be interpreted in light of the quality and representativeness of the available evidence.

This study also highlights at least three areas for future research efforts. First, expanding medicine quality surveillance, particularly to countries and regions that are under-sampled will help improve the precision of model estimates. It would also potentially enable more granular prevalence estimates, for example at a subnational level. Such surveillance should be undertaken using random sampling and include rural areas to help reduce bias in prevalence estimates. Second, linking these prevalence estimates to clinical outcomes could help quantify the public health burden of SF medicines. Third, adaptation of our modelling approach to other classes of medicines, such as antibiotics, could highlight broader patterns in the distribution of poor-quality medicines. Lastly, while our study identified country-specific temporal changes in the prevalence of substandard and falsified antimalarials, it was not designed to determine the factors driving these trends. Future studies should investigate the influence of changes in medicines regulatory systems, post-marketing surveillance, procurement practices, and other country-level interventions using longitudinal data to better understand the drivers of temporal changes in medicine quality.

SF antimalarials remain a pervasive yet unevenly distributed threat across Africa. While several countries have been able to achieve encouraging declines since the early 2000s, much of the continent still has high prevalence, especially in West and Central Africa. Achieving Sustainable Development Goal 3.8, which advocates for safe, effective, quality, and affordable medicines, will require sustained investment in regulatory capacity, cross*Ö*border collaboration, and transparent reporting of medicine quality data. The spatiotemporal estimates provided here offer a foundation for such efforts.

## CONTRIBUTORS

All authors contributed to the study. Study conceptualization: SC, JAM, LBA, BC, PN, CC, KVA, SMN, NS, PM. LBA was responsible for data processing and analysis. SC, JAM, and LBA did the modelling analysis. LBA created the visualizations. LBA drafted the manuscript, and all authors read it and contributed to its editing. We hereby confirm that all authors have full access to all the data in the study and agreed to submit for publication. SC, JAM, and LBA have directly accessed and verified the underlying data reported in the manuscript.

## Data Availability

All data are publicly available and code will be shared on GitHub.

https://github.com/babulawrence/SF-Antimalarials-in-Africa

## ACKNOWLEDGEMENTS

We would like to acknowledge the members of the FORESFA Collaboration for the diverse discussions and cooperation that benefitted this research.

## ETHICAL APPROVAL STATEMENT

This study is a secondary analysis of data from publicly available data on the quality of antimalarial medicines. The original publications should provide details regarding the ethical approval of the studies.

## AUTHORS’ ADDRESSES

Lawrence B. Adipo, Jessica A. Mendes, Ngan Thi Do, Kerlijn Van Assche, Stella Maris Nanyonga, Nicole Stoesser, Christiane Dolecek, Paul N. Newton, Ben S. Cooper, Celine Caillet and Sean M. Cavany, Nuffield Department of Medicine, University of Oxford, Oxford, United Kingdom. Paula Moraga, Division of Computer, Electrical and Mathematical Sciences and Engineering, King Abdullah University of Science and Technology, Thuwal, Saudi Arabia

