## Supplementary material for "The spatiotemporal distribution of substandard and falsified antimalarial medicines in Africa, 1996-2019"

**Supplementary information**

**S1, Supplementary Table 1: Description of the World Governance Indicators**

| Variable | Definition | Estimate |
|---|---|---|
| Voice and accountability | Reflects perceptions of the extent to which a country's citizens are able to participate in selecting their government, as well as freedom of expression, freedom of association, and a free media. | Estimate of governance (ranges from approximately -2.5 (weak) to 2.5 (strong) governance performance) |
| Political stability and absence of violence or terrorism | Political stability and absence of violence/terrorism measures perceptions of the likelihood of political instability and/or politically-motivated violence, including terrorism. | Estimate of governance (ranges from approximately -2.5 (weak) to 2.5 (strong) governance performance) |
| Government effectiveness | Reflects perceptions of the quality of public services, the quality of the civil service and the degree of its independence from political pressures, the quality of policy formulation and implementation, and the credibility of the government's commitment to such policies. | Estimate of governance (ranges from approximately -2.5 (weak) to 2.5 (strong) governance performance) |
| Regulatory quality | Reflects perceptions of the ability of the government to formulate and implement sound policies and regulations that permit and promote private sector development. | Estimate of governance (ranges from approximately -2.5 (weak) to 2.5 (strong) governance performance) |
| Rule of law | Reflects perceptions of the extent to which agents have confidence in and abide by the rules of society, and in particular the quality of contract enforcement, property rights, the police, and the courts, as well as the likelihood of crime and violence. | Estimate of governance (ranges from approximately -2.5 (weak) to 2.5 (strong) governance performance) |
| Control of corruption | Reflects perceptions of the extent to which public power is exercised for private gain, including both petty and grand forms of corruption, as well as "capture" of the state by elites and private interests. | Estimate of governance (ranges from approximately -2.5 (weak) to 2.5 (strong) governance performance) |

*Data source: Aggregate Governance Indicators 1996-2022. www.govindicators.org*
*The six aggregate indicators are based on over 30 underlying data sources reporting the perceptions of governance of a large number of survey respondents and expert assessments worldwide.  Details on the underlying data sources, the aggregation method, and the interpretation of the indicators can be found in the WGI methodology paper (source: Kaufmann D, Kraay A, Mastruzzi M. The worldwide governance indicators: Methodology and analytical issues1. Hague journal on the rule of law. 2011;3(2):220-46)*

**S2, Supplementary Table 2: Malaria incidence, population, and GDP per capita**

| Variable | Definition | Units |
|---|---|---|
| Malaria incidence[1] | Number of new malaria infections per 1000 people at risk | Per 1000 persons |
| Population[2] | Total population by country | Millions |
| GDP per capita[2] | GDP per capita is gross domestic product divided by midyear population. GDP is the sum of gross value added by all resident producers in the economy plus any product taxes and minus any subsidies not included in the value of the products. It is calculated without making deductions for depreciation of fabricated assets or for depletion and degradation of natural resources. Data are in current U.S. dollars. | US dollars |

*Data source [1]. Institute for Health Metrics and Evaluation, Global Burden of Disease (2024)*

*Data source [2]. World Bank. Word development indicators (2024)*

**S3, Supplementary Figure 1: Number of surveys, observed SF proportions and total number of samples**

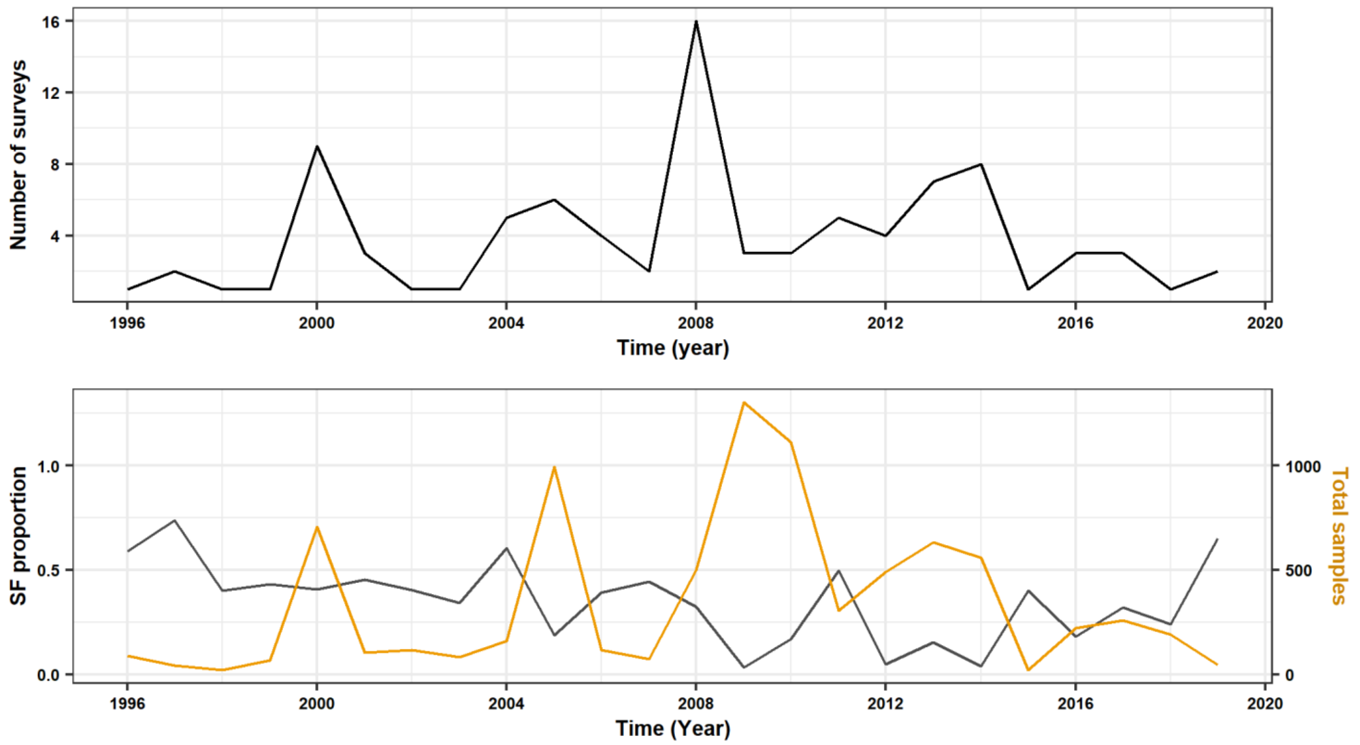


*Online supplemental figure 2: Number of surveys on quality of antimalarials (top) and observed SF proportions and total number of samples (bottom) plotted against the year of study, 1996-2019*

*Alt Text: Plots showing number of surveys by year (top) and SF proportions and total number of samples from observed data (bottom)*

**S4, Supplementary Table 3: Variance inflation factor (VIF) analysis of predictor variables**

| **Variable** | **VIF value** |
|---|---|
| Rule of law | 8.5 |
| Government effectiveness | 5.2 |
| Political stability | 5.0 |
| Control of corruption | 4.3 |
| Regulatory quality | 4.1 |
| Voice and accountability | 3.0 |
| Population density | 2.1 |
| GDP per capita | 1.4 |
| Malaria incidence | 1.2 |

*Supplementary Table 3: Variance inflation factor (VIF) analysis of predictor variables included in the Bayesian spatiotemporal model. The table presents the VIF values for each predictor variable used to assess multicollinearity. All VIF values were below the predefined threshold (<10), indicating no evidence of problematic multicollinearity.*

*Alt text: Table showing the variance inflation factor (VIF) values for each predictor variable included in the Bayesian spatiotemporal model. The table was used to assess multicollinearity among predictor variables before model fitting.*

**S5, Supplementary Figure 2: Correlation matrix of SF antimalarials and predictor variables**

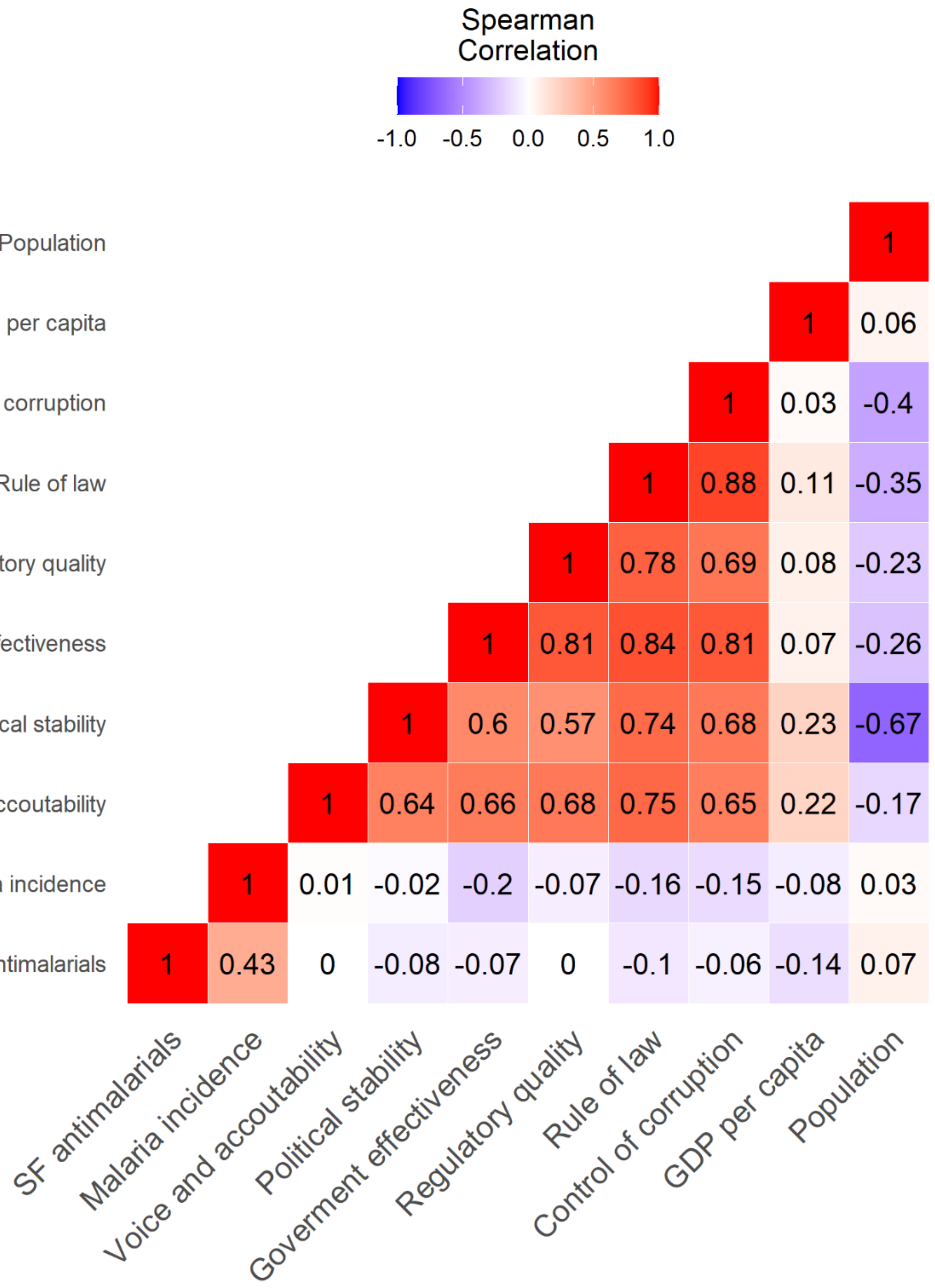


*Online supplemental figure 2: Correlation matrix of SF antimalarials and predictor variables showing the association between all variables with each other. Correlation coefficients of 0.90 to 1.00 (-0.90 to -1.00) are considered very high positive (negative) correlation; 0.70 to 0.90 (-0.70 to -0.90) are high positive (negative) correlation; 0.50 to 0.70 (-0.50 to -0.70) are moderate positive (negative) correlation; 0.30 to 0.50 (-0.30 to -0.50) are low positive (negative) correlation; 0.00 to 0.30 (0.00 to -0.30) are negligible correlation.*

*Alt Text: Plot showing correlation matrix of all variables with each other.*

**S6, Supplementary figure 3: Spatio-temporal maps showing 95% credibility of the predicted proportions from model 3**

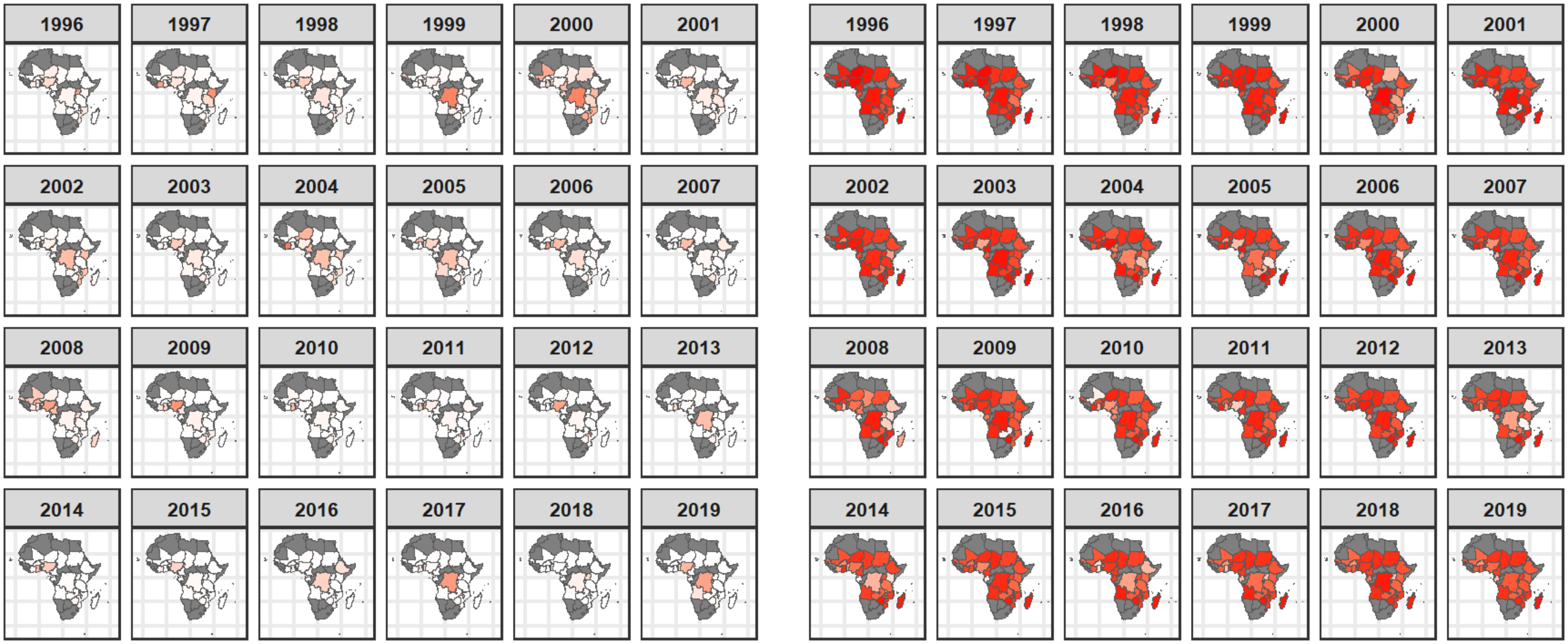


*Online supplemental figure 3: Spatio-temporal maps showing the 95% credible intervals of the predicted SF proportions (Figure 3) in Africa from 1996 to 2019. Estimates from Random walk and spatiotemporal interaction (model 3).*

*Alt text: Maps showing 95% credible intervals of the predicted proportions (Figure 3) of SF antimalarials in Africa from 1996 to 2019.*

**S7, Supplementary Figure 4: Time series of SF predictions from parametric (model 1, top) and non-parametric (model 2, bottom) models**

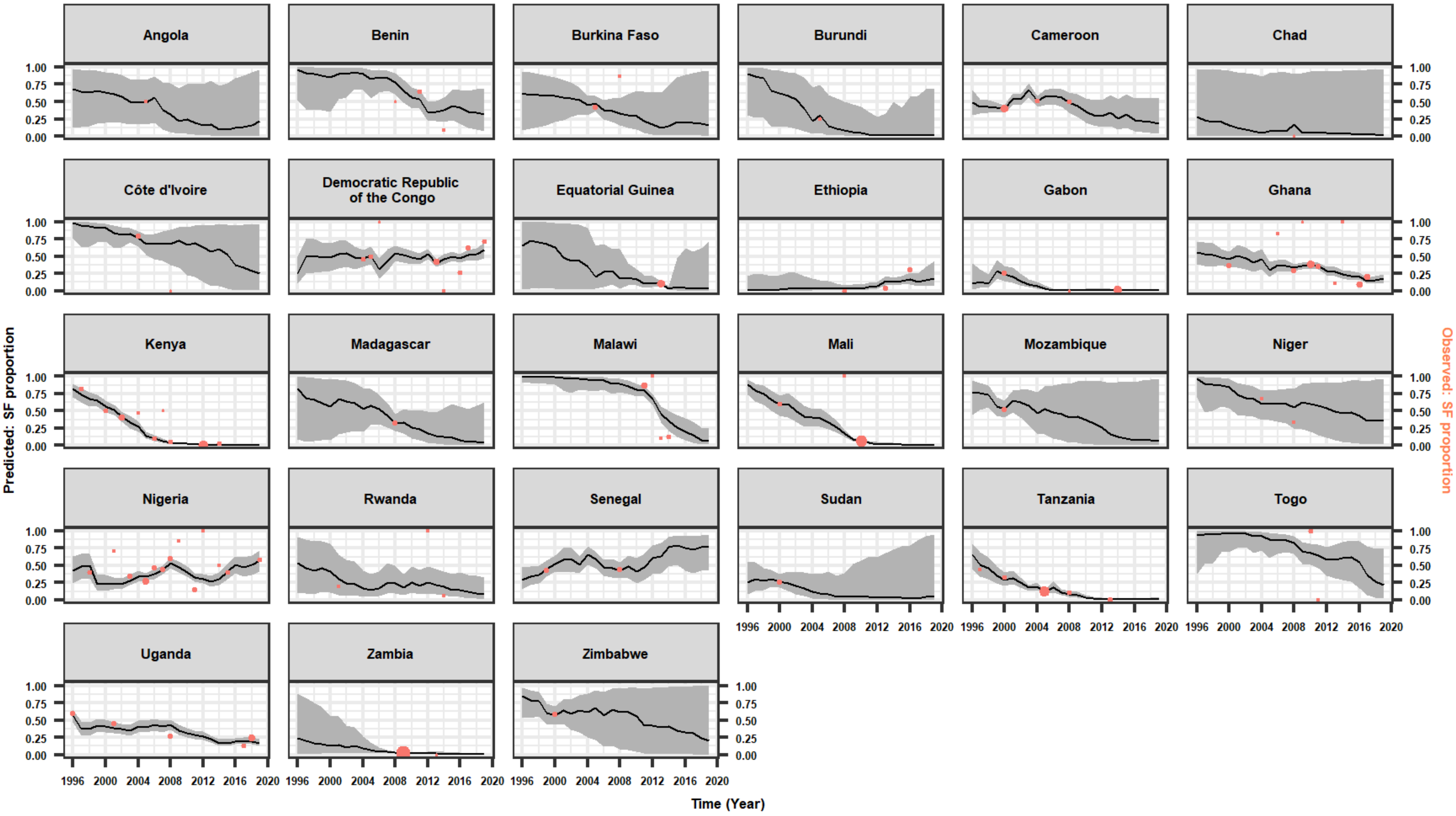


*Online supplemental figure 4: Annual trend of SF proportions in Africa from 1996 to 2019. Data in coral color represents observed proportions, predicted estimates denoted by the dark line and their 2.5% and 97.5% credible intervals in the grey region. Predictions from the parametric model*

*Alt text: Plots of predicted mean estimates and their 95% credible intervals showing the trends in SF proportions of antimalarials in Africa from 1996 to 2019. Predictions from the parametric model (1)*

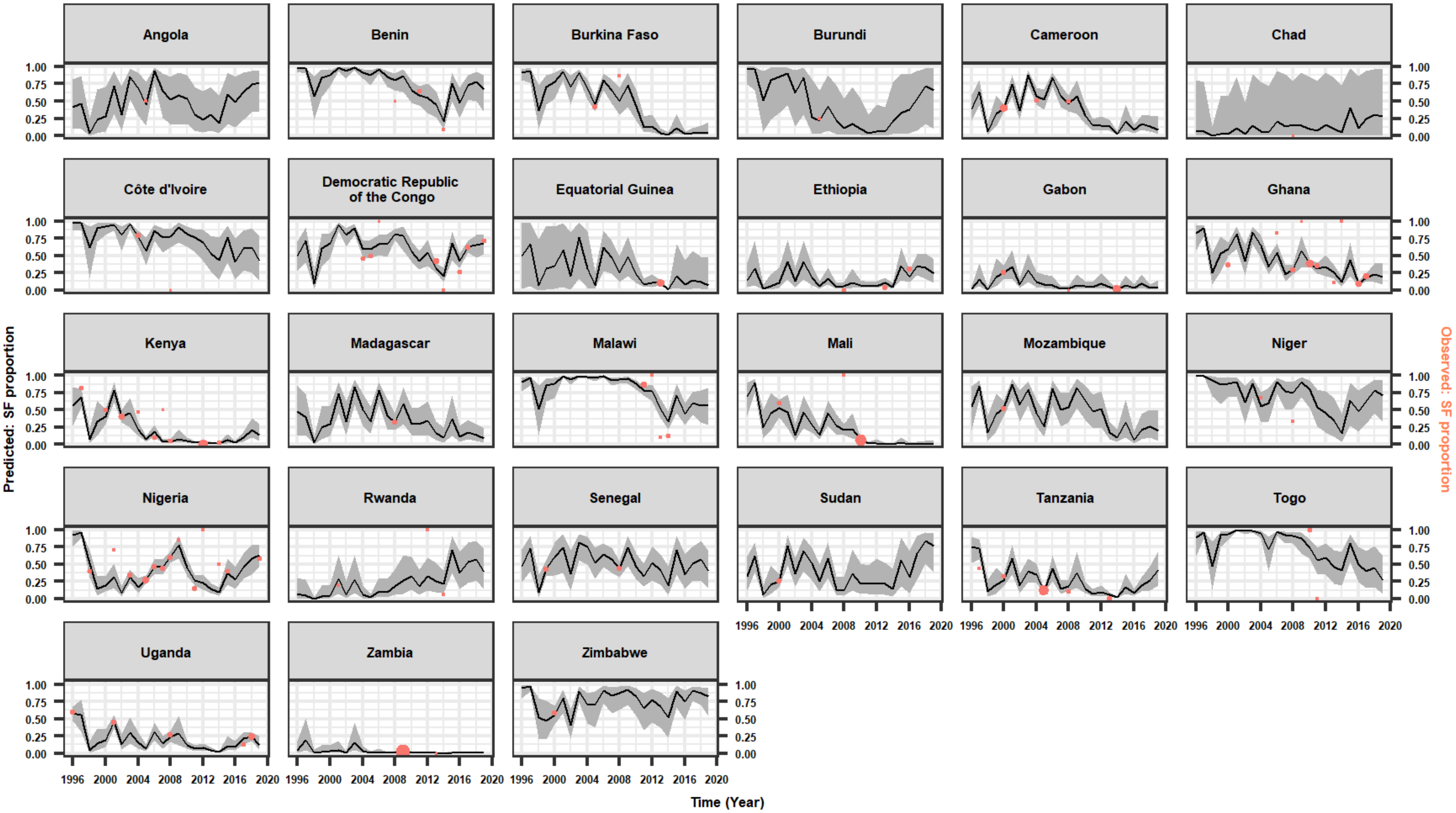


*Online supplemental figure 5: Annual trend of SF proportions in Africa from 1996 to 2019. Data in coral color represents observed proportions, predicted estimates denoted by the dark line and their 2.5% and 97.5% credible intervals in the grey region. Predictions from the nonparametric model.*

*Alt Text: Plots of predicted mean estimates and their 95% credible intervals showing the trends in SF proportions of antimalarials in Africa from 1996 to 2019. Predictions from the random walk (RW1) model (2).*

**S8, Supplementary Table 4: Raw data used to generate the prevalence estimates of SF antimalarials**

| Country | Year of survey | Total number of samples | Samples failing at least one test | Type of sampling | Testing Methodology | Source of data |
|---|---|---|---|---|---|---|
| Angola | 2005 | 6 | 3 | Convenience survey | High Performance Liquid Chromatography (HPLC) | 1* |
| Benin | 2008 | 2 | 1 | Random survey | UV Spectrophotometry | 2* |
| Benin | 2011 | 40 | 26 | Convenience survey | UV Spectrophotometry, Mass Spectrometry (MS) | 3 |
| Benin | 2014 | 12 | 1 | Convenience survey | Thin Layer Chromatography TLC, MS Mass spectrometry | 4* |
| Burkina Faso | 2005 | 77 | 32 | Convenience survey | UV Spectrophotometry, Visual inspection | 5 |
| Burkina Faso | 2008 | 7 | 6 | Random survey | UV Spectrophotometry | 2 |
| Burundi | 2005 | 4 | 1 | Convenience survey | High Performance Liquid Chromatography (HPLC) | 1 |
| Cameroon | 2000 | 283 | 112 | Convenience survey | Raman Spectroscopy, UV spectrophotometry | 6 |
| Cameroon | 2004 | 37 | 19 | Convenience survey | MS Mass spectrometry, High Performance Liquid Chromatography (HPLC) | 7* |
| Cameroon | 2008 | 32 | 16 | Convenience survey | High Performance Liquid Chromatography (HPLC) | 8* |
| Chad | 2008 | 1 | 0 | Random survey | UV Spectrophotometry | 2 |
| Democratic Republic of the Congo | 2004 | 7 | 1 | Convenience survey | High Performance Liquid Chromatography (HPLC), UV-Vis Spectrophotometry | 9* |
| Democratic Republic of the Congo | 2004 | 28 | 15 | Convenience survey | MS Mass spectrometry, High Performance Liquid Chromatography (HPLC) | 10 |
| Democratic Republic of the Congo | 2005 | 20 | 10 | Convenience survey | High Performance Liquid Chromatography (HPLC) | 1 |
| Democratic Republic of the Congo | 2006 | 1 | 1 | Convenience survey | MS Mass spectrometry, High Performance Liquid Chromatography (HPLC) | 11* |
| Democratic Republic of the Congo | 2013 | 150 | 63 | Convenience survey | MS Mass spectrometry, High Performance Liquid Chromatography (HPLC), UV-Vis Spectrophotometry | 12 |
| Democratic Republic of the Congo | 2014 | 9 | 0 | Convenience survey | Thin Layer Chromatography TLC, MS Mass spectrometry | 4 |
| Democratic Republic of the Congo | 2016 | 45 | 12 | Random survey | UV-Vis Spectrophotometry, High Performance Liquid Chromatography (HPLC), Raman Spectroscopy | 13 |
| Democratic Republic of the Congo | 2017 | 79 | 49 | Random survey | MS Mass spectrometry, High Performance Liquid Chromatography (HPLC) | 14 |
| Democratic Republic of the Congo | 2019 | 24 | 17 | Convenience survey | Raman Spectroscopy, Hardness test, Visual inspection, UV-Vis Spectrophotometry | 15 |
| Equitoria Guinea | 2013 | 309 | 31 | Random survey | UV-Vis Spectrophotometry, Microbiological assay | 16 |
| Ethiopia | 2008 | 25 | 0 | Convenience survey | High Performance Liquid Chromatography (HPLC) | 8 |

| | | | | | | |
|---|---|---|---|---|---|---|
| Ethiopia | 2013 | 74 | 2 | Convenience survey | Raman Spectroscopy, High Performance Liquid Chromatography (HPLC), Visual inspection | 17 |
| Ethiopia | 2016 | 60 | 18 | Random survey | High Performance Liquid Chromatography (HPLC), UV-Vis Spectrophotometry | 18 |
| Gabon | 2000 | 53 | 13 | Convenience survey | High Performance Liquid Chromatography (HPLC), UV-Vis Spectrophotometry | 19* |
| Gabon | 2008 | 2 | 0 | Random survey | UV Spectrophotometry | 2 |
| Gabon | 2014 | 432 | 3 | Random survey | UV Spectrophotometry , Raman Spectroscopy, High Performance Liquid Chromatography (HPLC) | 20 |
| Ghana | 2000 | 55 | 20 | Convenience survey | High Performance Liquid Chromatography (HPLC), UV-Vis Spectrophotometry | 19 |
| Ghana | 2006 | 17 | 14 | Convenience survey | Thin Layer Chromatography TLC, UV Spectrophotometry | 21 |
| Ghana | 2008 | 76 | 22 | Convenience survey | High Performance Liquid Chromatography (HPLC) | 8 |
| Ghana | 2009 | 2 | 2 | Convenience survey | MS Mass spectrometry, High Performance Liquid Chromatography (HPLC) | 11 |
| Ghana | 2010 | 17 | 16 | Convenience survey | Thin Layer Chromatography TLC, UV Spectrophotometry, Raman Spectroscopy | 22 |
| Ghana | 2010 | 9 | 2 | Convenience survey | Microbiological assay , UV Spectrophotometry , Raman | 23 |
| Ghana | 2010 | 254 | 90 | Random survey | Microbiological assay , UV Spectrophotometry , Raman | 24 |
| Ghana | 2011 | 65 | 15 | Convenience survey | Microbiological assay , UV Spectrophotometry , Thin Layer Chromatography TLC | 25* |
| Ghana | 2011 | 16 | 13 | Random survey | Microbiological assay , UV Spectrophotometry , Thin Layer Chromatography TLC | 26 |
| Ghana | 2013 | 10 | 1 | Random survey | Microbiological assay , UV Spectrophotometry , Thin Layer Chromatography TLC | 27 |
| Ghana | 2014 | 4 | 4 | Random survey | PH measurement, Microbiological assay, Microbiological assay , UV Spectrophotometry | 28 |
| Ghana | 2016 | 90 | 4 | Random survey | Microbiological assay | 29 |
| Ghana | 2016 | 25 | 6 | Convenience survey | High Performance Liquid Chromatography (HPLC), MS Mass spectrometry , UV Spectrophotometry | 30 |
| Ghana | 2017 | 150 | 30 | Random survey | High Performance Liquid Chromatography (HPLC), MS Mass spectrometry , UV Spectrophotometry , Raman Spectroscopy | 31 |
| Ivory Coast | 2004 | 56 | 44 | Random survey | Visual inspection, CD-3 ultraviolet | 32 |
| Ivory Coast | 2008 | 1 | 0 | Random survey | UV Spectrophotometry | 2 |
| Kenya | 1997 | 33 | 27 | Convenience survey | Dissolution test, High Performance Liquid Chromatography (HPLC), UV Spectrophotometry | 33 |
| Kenya | 2000 | 44 | 22 | Convenience survey | High Performance Liquid Chromatography (HPLC), UV-Vis Spectrophotometry | 19 |
| Kenya | 2002 | 116 | 47 | Convenience survey | Dissolution test, High Performance Liquid Chromatography (HPLC), UV Spectrophotometry , Microbiological assay , Raman | 34 |
| Kenya | 2004 | 17 | 8 | Convenience survey | High Performance Liquid Chromatography (HPLC) UV-Vis Spectrophotometry | 9 |

| | | | | | | |
|---|---|---|---|---|---|---|
| Kenya | 2006 | 41 | 4 | Convenience survey | High Performance Liquid Chromatography (HPLC), Raman, UV spectrophotometry, Dissolution test | 35 |
| Kenya | 2007 | 2 | 1 | Convenience survey | MS Mass spectrometry, High Performance Liquid Chromatography (HPLC) | 11 |
| Kenya | 2008 | 44 | 2 | Convenience survey | High Performance Liquid Chromatography (HPLC) | 8 |
| Kenya | 2012 | 471 | 2 | Convenience survey | Haskins test, Raman Spectroscopy | 36 |
| Kenya | 2014 | 39 | 0 | Convenience survey | High Performance Liquid Chromatography (HPLC), Microbiological assay , UV spectrophotometry , Dissolution test | 37 |
| Kenya | 2014 | 1 | 1 | Convenience survey | Analytical methodology not reported; High Performance Liquid Chromatography (HPLC) is suggested, most likely assay technique based on quality control practice | 38 |
| Madagascar | 2008 | 53 | 17 | Convenience survey | Raman Spectroscopy, Infrared (IR) spectroscopy, High Performance Liquid Chromatography (HPLC), UV spectrophotometry | 39* |
| Malawi | 2011 | 112 | 99 | Convenience survey | Microbiological test, UV spectrophotometry, Visual inspection, Raman Spectroscopy | 40 |
| Malawi | 2013 | 10 | 1 | Random survey | Thin Layer Chromatography TLC, Visual inspection, UV spectrophotometry | 41 |
| Malawi | 2014 | 31 | 4 | Random survey | Disintegration test, Dissolution test, UV-Vis Spectrophotometry, High Performance Liquid Chromatography (HPLC) | 42 |
| Mali | 2000 | 40 | 24 | Convenience survey | High Performance Liquid Chromatography (HPLC), UV-Vis Spectrophotometry | 19 |
| Mali | 2008 | 3 | 3 | Random survey | UV Spectrophotometry | 2 |
| Mali | 2010 | 810 | 57 | Convenience survey | Microbiological test, Disintegration test, Thin Layer Chromatography TLC, UV spectrophotometry | 43 |
| Mozambique | 2000 | 63 | 33 | Convenience survey | High Performance Liquid Chromatography (HPLC), UV-Vis Spectrophotometry | 19 |
| Niger | 2004 | 15 | 10 | Convenience survey | MS Mass spectrometry, High Performance Liquid Chromatography (HPLC) | 7 |
| Niger | 2008 | 3 | 1 | Random survey | UV Spectrophotometry | 2 |
| Nigeria | 1998 | 20 | 8 | Convenience survey | UV spectrophotometry, Microbiological assay, Raman | 44 |
| Nigeria | 2001 | 7 | 5 | Convenience survey | Disintegration test, Friability test, Dissolution test, hardness test, High Performance Liquid Chromatography (HPLC), UV spectrophotometry | 45 |
| Nigeria | 2003 | 32 | 9 | Convenience survey | Disintegration test, Dissolution test, High Performance Liquid Chromatography (HPLC), UV spectrophotometry | 46 |
| Nigeria | 2003 | 50 | 19 | Convenience survey | UV spectrophotometry, Thin Layer Chromatography TLC, Visual inspection, Microbiological assay, Raman | 47 |
| Nigeria | 2005 | 225 | 60 | Random survey | High Performance Liquid Chromatography (HPLC), Dissolution test, UV spectrophotometry, Raman | 48 |
| Nigeria | 2006 | 18 | 8 | Convenience survey | Disintegration test, Dissolution test | 49 |
| Nigeria | 2006 | 9 | 5 | Convenience survey | UV spectrophotometry | 50 |
| Nigeria | 2006 | 9 | 1 | Convenience survey | Dissolution test, High Performance Liquid Chromatography (HPLC), Disintegration test, UV spectrophotometry | 51 |

| | | | | | | |
|---|---|---|---|---|---|---|
| Nigeria | 2006 | 20 | 12 | Random survey | Visual inspection, Disintegration test, Friability test, Dissolution test, UV spectrophotometry, Microbiological assay | 52 |
| Nigeria | 2007 | 70 | 31 | Convenience survey | High Performance Liquid Chromatography (HPLC), Disintegration test, Dissolution test, Visual inspection, Microbiological assay, UV spectrophotometry | 53 |
| Nigeria | 2008 | 64 | 37 | Convenience survey | High Performance Liquid Chromatography (HPLC) | 8 |
| Nigeria | 2008 | 3 | 3 | | High Performance Liquid Chromatography (HPLC) , Microbiological assay ,Disintegration test, Hardness test, UV-Vis Spectrophotometry | 54 |
| Nigeria | 2009 | 13 | 11 | Convenience survey | MS Mass spectrometry, High Performance Liquid Chromatography (HPLC) , Raman, Microbiological assay | 55 |
| Nigeria | 2011 | 75 | 11 | Convenience survey | Microbiological assay, UV Spectrophotometry, Thin Layer Chromatography TLC | 25 |
| Nigeria | 2012 | 6 | 6 | Convenience survey | High Performance Liquid Chromatography (HPLC) | 56 |
| Nigeria | 2014 | 16 | 8 | Convenience survey | High Performance Liquid Chromatography (HPLC), Dissolution test, Mass uniformity test, Microbiological assay, Raman Spectroscopy, Infrared (IR) spectroscopy | 57 |
| Nigeria | 2015 | 20 | 8 | Convenience survey | Disintegration test, Hardness test, Friability test | 58 |
| Nigeria | 2019 | 22 | 13 | Convenience survey | Infrared (IR) spectroscopy, High Performance Liquid Chromatography (HPLC), Visual inspection, Raman Spectroscopy | 59 |
| Rwanda | 2001 | 5 | 1 | Convenience survey | High Performance Liquid Chromatography (HPLC), Dissolution test, UV spectrophotometry, Microbiological assay | 60 |
| Rwanda | 2012 | 3 | 3 | Convenience survey | Microbiological assay, Disintegration test, Friability test, Uniformity of mass, UV spectrophotometry | 61 |
| Rwanda | 2014 | 16 | 1 | Random survey | High Performance Liquid Chromatography (HPLC), MS Mass spectrometry | 62 |
| Senegal | 1999 | 67 | 29 | Convenience survey | Raman Spectroscopy, UV spectrophotometry, Microbiological assay | 63 |
| Senegal | 2008 | 63 | 28 | Convenience survey | Raman Spectroscopy, Infrared (IR) spectroscopy, High Performance Liquid Chromatography (HPLC), UV spectrophotometry | 39 |
| Sudan | 2000 | 76 | 20 | Convenience survey | High Performance Liquid Chromatography (HPLC), UV-Vis Spectrophotometry | 19 |
| Tanzania | 1997 | 9 | 4 | Convenience survey | High Performance Liquid Chromatography (HPLC), Dissolution test, UV-Vis Spectrophotometry | 64 |
| Tanzania | 2000 | 33 | 12 | Convenience survey | High Performance Liquid Chromatography (HPLC), Dissolution test, UV spectrophotometry, Microbiological assay | 65 |
| Tanzania | 2000 | 9 | 2 | Convenience survey | High Performance Liquid Chromatography (HPLC), Dissolution test, UV spectrophotometry, Microbiological assay, Thin Layer Chromatography | 66 |
| Tanzania | 2005 | 361 | 41 | Convenience survey | Dissolution test, UV spectrophotometry, High Performance Liquid Chromatography (HPLC), Raman Spectroscopy, Infrared (IR) spectroscopy | 67 |
| Tanzania | 2005 | 301 | 38 | Random survey | High Performance Liquid Chromatography (HPLC), MS Mass spectrometry, Raman | 68 |

| | | | | | | |
|---|---|---|---|---|---|---|
| Tanzania | 2008 | 38 | 4 | Convenience survey | High Performance Liquid Chromatography (HPLC) | 8 |
| Tanzania | 2013 | 80 | 0 | Random survey | Visual inspection, Disintegration test | 69 |
| Togo | 2010 | 22 | 22 | Convenience survey | Thin Layer Chromatography TLC | 70 |
| Togo | 2011 | 10 | 0 | Convenience survey | Microbiological assay, UV Spectrophotometry, Thin Layer Chromatography TLC | 25 |
| Uganda | 1996 | 88 | 52 | Random survey | Thin Layer Chromatography TLC, UV spectrophotometry, Microbiological assay, Raman | 71 |
| Uganda | 2001 | 92 | 41 | Convenience survey | High Performance Liquid Chromatography (HPLC), Microbiological assay, MS Mass spectrometry | 72 |
| Uganda | 2008 | 82 | 21 | Convenience survey | Raman Spectroscopy, Infrared (IR) spectroscopy, High Performance Liquid Chromatography (HPLC), UV spectrophotometry | 39 |
| Uganda | 2017 | 30 | 4 | Random survey | Thin Layer Chromatography TLC, UV spectrophotometry, Mass uniformity test, Microbiological test | 73 |
| Uganda | 2018 | 192 | 46 | Random survey | Thin Layer Chromatography TLC, Disintegration test, Dissolution test, Visual inspection | 74 |
| Zambia | 2009 | 1290 | 30 | Random survey | Thin Layer Chromatography TLC, Visual inspection, UV spectrophotometry, Raman Spectroscopy | 75 |
| Zambia | 2013 | 1 | 0 | Convenience survey | Raman Spectroscopy, Thin Layer Chromatography TLC, Raman Spectroscopy | 76 |
| Zimbabwe | 2000 | 52 | 30 | Convenience survey | High Performance Liquid Chromatography (HPLC), UV-Vis Spectrophotometry | 19 |

*Surveys conducted in more than one country.

*Table showing characteristics of all medicine quality surveys included in the Bayesian spatiotemporal analysis. The table summarises the country, year of data collection, sample size, sampling strategy, testing methodology, and the number of substandard and falsified antimalarial medicines identified in each survey.*

*Alt text: Table summarising the characteristics of all medicine quality surveys included in the study, including country, year of data collection, sample size, sampling strategy, testing methodology, and the number of substandard and falsified antimalarial medicines identified.*

**S9, Supplementary Table 5: Comparison of spatiotemporal models using different covariate combinations**

| **Model 1: Parametric time trend** | | | | |
|---|---|---|---|---|
| **Set of covariates** | **Covariates** | **DIC** | **WAIC** | **MLL** |
| 0 | Simple | 690.12 | 779.07 | -389.56 |
| 1 | Malaria incidence | 691.07 | 778.07 | -382.46 |
| 1 | Population | 681.39 | 763.59 | -396.45 |
| 1 | GDP per capita | 687.46 | 775.68 | -394.40 |
| 2 | Malaria incidence + GDP per capita | 687.41 | 774.96 | -387.54 |
| 2 | Malaria incidence + Population | 682.74 | 768.44 | -389.46 |
| 2 | GDP per capita + Population | 679.54 | 761.08 | -401.13 |
| 3 | Malaria incidence + GDP per capita + Population | 677.39 | 761.23 | -394.40 |
| 6 | Government effectiveness + Regulatory quality + Rule of law + Control of corruption + Voice and accountability + Political stability | 665.62 | 752.76 | -395.41 |
| 7 | Government effectiveness + Regulatory quality + Rule of law + Control of corruption + Voice and accountability + Political stability + Malaria incidence | 669.66 | 757.23 | -393.57 |
| 7 | Government effectiveness + Regulatory quality + Rule of law + Control of corruption + Voice and accountability + Political stability + GDP per capita | 666.15 | 755.39 | -400.57 |
| 7 | Government effectiveness + Regulatory quality + Rule of law + Control of corruption + Voice and accountability + Political stability + Population | 660.26 | 745.01 | -402.11 |
| 9 | Government effectiveness + Regulatory quality + Rule of law + Control of corruption + Voice and accountability + Political stability + Malaria incidence + GDP per capita + Population | 666.22 | 754.25 | -404.43 |
| **Model 2: Random walk** | | | | |
| **Set of covariates** | **Covariates** | **DIC** | **WAIC** | **MLL** |
| 0 | Simple | 796.69 | 965.27 | -434.31 |
| 1 | Malaria incidence | 776.41 | 929.94 | -418.66 |
| 1 | Population | 778.59 | 929.22 | -429.15 |
| 1 | GDP per capita | 797.44 | 966.57 | -440.44 |
| 2 | Malaria incidence + GDP per capita | 776.93 | 936.37 | -423.96 |
| 2 | Malaria incidence + Population | 759.34 | 902.59 | -423.62 |
| 2 | GDP per capita + Population | 775.53 | 926.83 | -434.37 |
| 3 | Malaria incidence + GDP per capita + Population | 759.93 | 904.80 | -429.66 |
| 6 | Government effectiveness + Regulatory quality + Rule of law + Control of corruption + Voice and accountability + Political stability | 656.54 | 744.16 | -398.22 |

| | | | | |
|---|---|---|---|---|
| 7 | Government effectiveness + Regulatory quality + Rule of law + Control of corruption + Voice and accountability + Political stability + Malaria incidence | 660.79 | 751.99 | -397.99 |
| 7 | Government effectiveness + Regulatory quality + Rule of law + Control of corruption + Voice and accountability + Political stability + GDP per capita | 659.60 | 749.54 | -403.17 |
| 7 | Government effectiveness + Regulatory quality + Rule of law + Control of corruption + Voice and accountability + Political stability + Population | 646.79 | 733.50 | -403.03 |
| 9 | Government effectiveness + Regulatory quality + Rule of law + Control of corruption + Voice and accountability + Political stability + Malaria incidence + GDP per capita + Population | 653.15 | 746.25 | -405.63 |
| **Model 3: Random walk + spatiotemporal interaction** | | | | |
| **Set of covariates** | **Covariates** | **DIC** | **WAIC** | **MLL** |
| 0 | Simple | 489.43 | 504.64 | -302.84 |
| 1 | Malaria incidence | 491.85 | 508.79 | -304.27 |
| 1 | Population | 488.88 | 503.50 | -310.89 |
| 1 | GDP per capita | 489.25 | 505.76 | -304.79 |
| 2 | Malaria incidence + GDP per capita | 491.95 | 510.80 | -306.03 |
| 2 | Malaria incidence + Population | 491.16 | 507.79 | -312.04 |
| 2 | GDP per capita + Population | 487.05 | 501.35 | -315.35 |
| 3 | Malaria incidence + GDP per capita + Population | 490.65 | 507.18 | -316.54 |
| 6 | Government effectiveness + Regulatory quality + Rule of law + Control of corruption + Voice and accountability + Political stability | 494.70 | 513.25 | -319.70 |
| 7 | Government effectiveness + Regulatory quality + Rule of law + Control of corruption + Voice and accountability + Political stability + Malaria incidence | 491.28 | 507.58 | -322.25 |
| 7 | Government effectiveness + Regulatory quality + Rule of law + Control of corruption + Voice and accountability + Political stability + GDP per capita | 493.82 | 512.48 | -323.27 |
| 7 | Government effectiveness + Regulatory quality + Rule of law + Control of corruption + Voice and accountability + Political stability + Population | 489.17 | 504.26 | -332.29 |
| 9 | Government effectiveness + Regulatory quality + Rule of law + Control of corruption + Voice and accountability + Political stability + Malaria incidence + GDP per capita + Population | 486.28 | 499.43 | -332.27 |

*Comparison of three Bayesian spatiotemporal models fitted using different combinations of covariates. Model performance was evaluated using the Deviance Information Criterion (DIC), Watanabe-Akaike Information Criterion (WAIC), and Marginal Log-Likelihood (MLL). Lower DIC and WAIC values indicate improved model fit, while higher MLL values indicate better model performance.*

*Alt text: Table comparing three Bayesian spatiotemporal models with different covariate combinations using DIC, WAIC, and MLL as model performance metrics.*

**S10, Supplementary Figure 6: Number of medicine quality surveys conducted each year by sampling strategy**

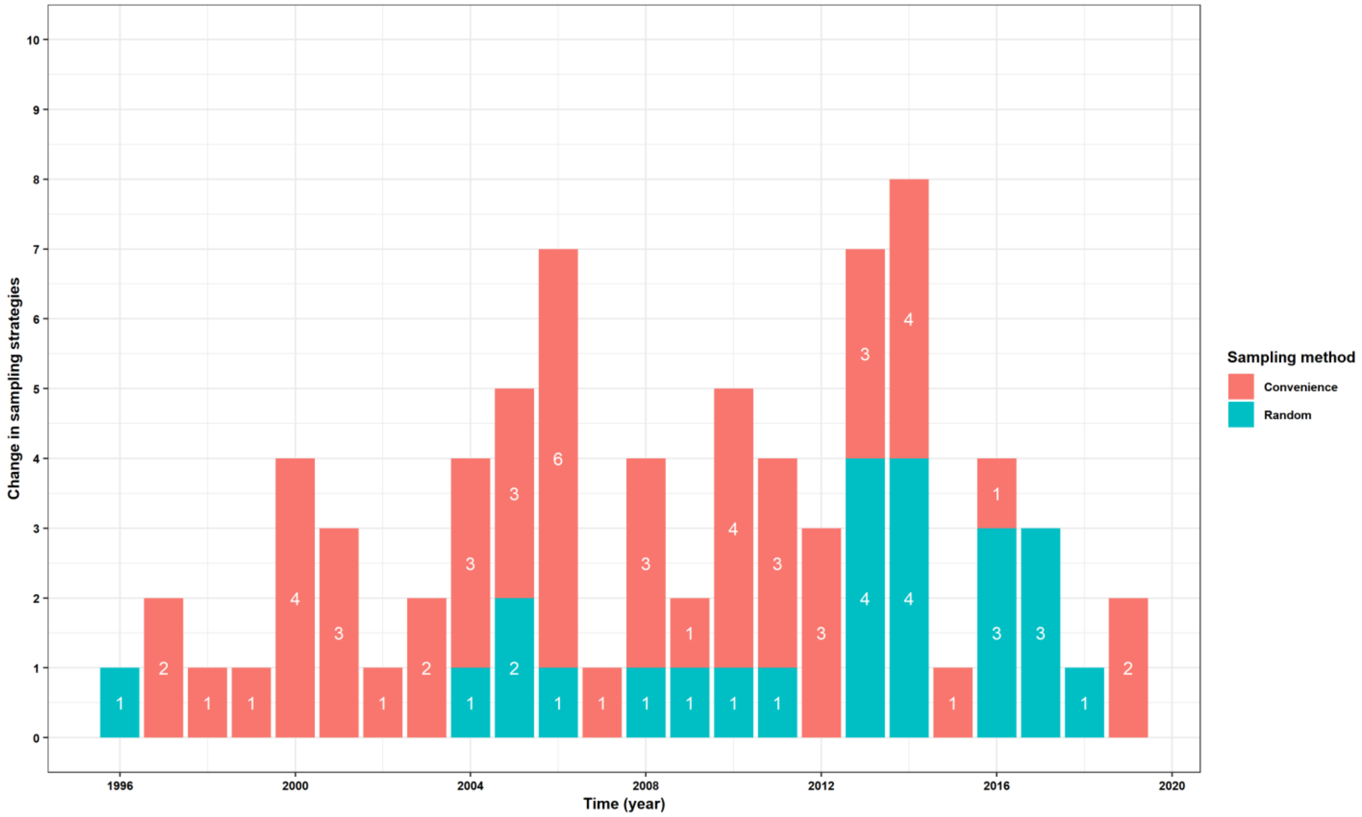


*Supplementary Figure 9: Annual distribution of medicine quality surveys included in the analysis according to sampling strategy. Bars represent the number of surveys conducted each year, stratified by convenience sampling and random sampling. The figure illustrates changes in survey methodologies over time, including the increasing use of random sampling in more recent years.*

*Alt text: Bar chart showing the annual number of medicine quality surveys conducted between 1996 and 2019, grouped by sampling strategy. Convenience sampling predominates in earlier years, while random sampling becomes more frequent in later years, although convenience sampling remains common throughout the study period.*

**S11, Supplementary Table 6. Sensitivity analysis excluding surveys with fewer than 11 tested medicine samples**

| Analysis | Surveys included | DIC | WAIC | Interpretation |
|---|---|---|---|---|
| Primary analysis | 76 | 486.28 | 499.43 | Baseline |
| Sensitivity analysis (Excluding surveys with < 11 samples) | 67 | 442.36 | 440.13 | Similar spatiotemporal patterns with improved model fit |

*The number of studies with fewer than 11 samples was nine.*

*This table compares the primary Bayesian spatiotemporal model with a sensitivity analysis excluding surveys that tested fewer than 11 medicine samples. It reports the number of surveys included, model fit statistics (DIC and WAIC), and shows that excluding very small surveys improved model fit while producing similar spatiotemporal prevalence estimates.*

*Alt text: Sensitivity analysis excluding surveys with fewer than 11 tested medicine samples and comparison of model performance with the primary analysis.*

### Source of Data


1. Gaudiano MC, Di Maggio A, Cocchieri E, Antoniella E, Bertocchi P, Alimonti S, Valvo L, 2007. Medicines informal market in Congo, Burundi and Angola: counterfeit and sub-standard antimalarials. *Malar J 6:* 22.
2. Sawadogo C, Amood Al-Kamarany M, Al-Mekhlafi H, Elkarbane M, Al-Adhroey A, Cherrah Y, Bouklouze A, 2011. Quality of chloroquine tablets available in Africa. *Ann Trop Med Parasitol 105:* 447-453.
3. Baba-Moussa F, Bonou J, Ahouandjinou H, Dougnon TV, Kpavode L, Raimi N, Ogouyemi-Hounto A, Abdoulaye I, Toukourou F, Loko F, 2015. Quality control of selected antimalarials sold in the illicit market: an investigation conducted in Porto-Novo City (Republic of Benin). *Adv Biosci Biotechnol 6:* 637-644.
4. Yemoa A, Habyalimana V, K. Mbinze J, Crickboom V, Muhigirwa B, Ngoya A, Sacre P-Y, Gbaguidi F, Quetin-Leclercq J, Hubert P, 2017. Detection of poor quality artemisinin-based combination therapy (ACT) medicines marketed in Benin using simple and advanced analytical techniques. *Current Drug Safety 12:* 178-186.
5. Tipke M, Diallo S, Coulibaly B, Störzinger D, Hoppe-Tichy T, Sie A, Müller O, 2008. Substandard anti-malarial drugs in Burkina Faso. *Malar J 7:* 95.
6. Basco L, 2004. Molecular epidemiology of malaria in Cameroon: 19. Quality of antimalarial drugs used for self-medication. *Am J Trop Med Hyg 70*.
7. Pouillot R, Bilong C, Boisier P, Ciss M, Moumouni A, Amani I, Nabeth P, 2008. Le circuit informel des médicaments à Yaoundé et à Niamey: étude de la population des vendeurs et de la qualité des médicaments distribués. *Bull Soc Pathol Exot 101:* 113-118.
8. Organization WH, 2011. Survey of the quality of selected antimalarial medicines circulating in six countries of sub-Saharan Africa. *World Health Organization 118*.
9. Atemnkeng MA, De Cock K, Plaizier-Vercammen J, 2007. Quality control of active ingredients in artemisinin-derivative antimalarials within Kenya and DR Congo. *Trop Med Int Health 12:* 68-74.
10. Atemnkeng M, Chimanuka B, Plaizier-Vercammen J, 2007. Quality evaluation of chloroquine, quinine, sulfadoxine–pyrimethamine and proguanil formulations sold on the market in East Congo DR. *J Clin Pharm Ther 32:* 123-132.
11. Newton PN, Green MD, Mildenhall DC, Plançon A, Nettey H, Nyadong L, Hostetler DM, Swamidoss I, Harris GA, Powell K, 2011. Poor quality vital anti-malarials in Africa-an urgent neglected public health priority. *Malar J 10:* 352.
12. Mufusama JP, Ndjoko Ioset K, Feineis D, Hoellein L, Holzgrabe U, Bringmann G, 2018. Quality of the antimalarial medicine artemether–lumefantrine in 8 cities of the Democratic Republic of the Congo. *Drug Testing and Analysis 10:* 1599-1606.
13. Mavungu Landu DJ, Frédérich M, Manzambi Kuwekita J, Bongo-Pasi Nswe C, Mbinze J, Liégeois S, Kalenda Tshilombo N, Kwete Minga M, Ciza Hamuli P, Hubert P, 2020. Quality of antimalarials in Kinshasa peri-urban areas with regard to local pharmaceutical legislation and regulation. *International Health 12:* 253-263.
14. Schiavetti B, Wynendaele E, De Spiegeleer B, Mbinze GJ, Kalenda N, Marini R, Melotte V, Hasker E, Meessen B, Ravinetto R, 2018. The quality of medicines used in children and supplied by private pharmaceutical wholesalers in Kinshasa, Democratic Republic of Congo: a prospective survey. *The American journal of tropical medicine and hygiene 98:* 894.
15. Bukavu D, Pharmaceutical Quality Of Antimalarial Drugs: Quinine Sulfate And Artemether/Lumefantrine Tablets Sold On Bukavu Market.
16. Kaur H, Allan EL, Mamadu I, Hall Z, Green MD, Swamidos I, Dwivedi P, Culzoni MJ, Fernandez FM, Garcia G, 2017. Prevalence of substandard and falsified artemisinin-based combination antimalarial medicines on Bioko Island, Equatorial Guinea. *BMJ global health 2*.

17. Belew S, Suleman S, Mohammed T, Mekonnen Y, Duguma M, Teshome H, Bayisa B, Wynendaele E, D'Hondt M, Duchateau L, 2019. Quality of fixed dose artemether/lumefantrine products in Jimma Zone, Ethiopia. *Malar J 18:* 236.
18. Abuye H, Abraham W, Kebede S, Tatiparthi R, Suleman S, 2020. Physicochemical quality assessment of antimalarial medicines: chloroquine phosphate and quinine sulfate tablets from drug retail outlets of South-West Ethiopia. *Infection and drug resistance:* 691-701.
19. Maponga C, Ondari C, 2003. The quality of antimalarials: A study in selected African countries. *The quality of antimalarials: a study in selected african countries*, x, 54-x, 54.
20. Visser BJ, Meerveld-Gerrits J, Kroon D, Mougoula J, Vingerling R, Bache E, Boersma J, van Vugt M, Agnandji ST, Kaur H, 2015. Assessing the quality of anti-malarial drugs from Gabonese pharmacies using the MiniLab®: a field study. *Malar J 14:* 273.
21. Ofori-Kwakye K, Asantewaa Y, Gaye O, 2009. Quality of artesunate tablets sold in pharmacies in Kumasi, Ghana.
22. El-Duah M, Ofori-Kwakye K, 2012. Substandard artemisinin-based antimalarial medicines in licensed retail pharmaceutical outlets in Ghana. *J Vector Borne Dis 49:* 131-139.
23. Nyarko EA, Nettey H, 2013. Quality assessment of artemether/lumefantrine tablets sampled from pharmacies in Accra, using the MVHimagePCv8. exe color software. *Pharmacology & Pharmacy 4:* 567-572.
24. Tivura M, Asante I, van Wyk A, Gyaase S, Malik N, Mahama E, Hostetler DM, Fernandez FM, Asante KP, Kaur H, 2016. Quality of artemisinin-based combination therapy for malaria found in Ghanaian markets and public health implications of their use. *BMC Pharmacology and Toxicology 17:* 48.
25. Bate R, Hess K, Tren R, Mooney L, Cudjoe F, Ayodele T, Attaran A, 2012. Subsidizing artemisinin-based combination therapies: a preliminary investigation of the Affordable Medicines Facility–malaria. *Res Rep Trop Med:* 63-68.
26. Affum AO, Lowor S, Osae SD, Dickson A, Gyan BA, Tulasi D, 2013. A pilot study on quality of artesunate and amodiaquine tablets used in the fishing community of Tema, Ghana. *Malar J 12:* 220.
27. Prah J, Ameyaw EO, Afoakwah R, Fiawoyife P, Oppong-Danquah E, Boampong JN, 2016. Quality Assessment of Artemether-Lumefantrine Samples and Artemether Injections Sold in the Cape Coast Metropolis. *J Trop Med 2016:* 8602619.
28. Frimpong G, Ofori-Kwakye K, Kuntworbe N, Buabeng KO, Osei YA, Boakye-Gyasi ME, Adi-Dako O, 2018. Quality assessment of some essential children's medicines sold in licensed outlets in Ashanti Region, Ghana. *J Trop Med 2018:* 1494957.
29. Opoku S, Nyanor I, 2019. Qualitative and Quantitative Microbiological Studies of Paediatric Artemether-Lumefantrine Dry Powders and Paracetamol Syrups Obtained from Selected Drug Stores in Accra, Ghana. *J Trop Med 2019:* 7062016.
30. Opuni KF-M, Nettey H, Larbi MA, Amartey SNA, Nti G, Dzidonu A, Owusu-Danso P, Owusu NA, Nyarko AK, 2019. Usefulness of combined screening methods for rapid detection of falsified and/or substandard medicines in the absence of a confirmatory method. *Malar J 18:* 403.
31. Boakye-Agyeman F, 2017. Quantifying the Quality of Antimalarial Drugs in Ghana.
32. SONHON BT, 2012. Les médicaments de contrebande à base d'amoxicilline commercialisés à Abidjan (Cote d'Ivoire): Enquête et contrôle de qualité.
33. Kibwage IO, Ngugi J, 2000. Sulphadoxine/pyrimethamine tablet products on the Kenyan market: quality concerns. *The East and Central African Journal of Pharmaceutical Sciences 3:* 14-19.
34. Amin A, Snow R, Kokwaro G, 2005. The quality of sulphadoxine-pyrimethamine and amodiaquine products in the Kenyan retail sector. *J Clin Pharm Ther 30:* 559-565.

35. The Pharmacy & Poisons Board K, 2007. *Antimalarial Medicines in Kenya, Availability, Quality and Registration status*. Available at: http://apps.who.int/medicinedocs/en/d/Js16424e/. Accessed.
36. Division of Malaria Control and Pharmacy and Poisons Board, 2013. *Monitoring the Quality of Antimalarial Medicines Circulating in Kenya (2013)*. Available at: http://pharmacyboardkenya.org/search/results?q=Monitoring%20the%20Quality%20of%20Antimalarial%20Medicines%20Circulating%20in%20Kenya(2013). Accessed.
37. Ndwigah S, Stergachis A, Abuga K, Mugo H, Kibwage I, 2018. The quality of anti-malarial medicines in Embu County, Kenya. *Malar J 17:* 330.
38. Daily Nation, 2017. *Malaria drug Duo-Cotecxin to be recalled from Kenyan market*. Available at: https://www.nation.co.ke/news/Malaria-drug-Duo-Cotecxin-recalled-Kenya/1056-4227416-dd6okaz/index.html; https://www.businessdailyafrica.com/news/Sub-standard-malaria-drug-set-for-recall/539546-4227582-13b02y7/index.html. Accessed.
39. Organization WH, 2009. Survey of the quality of selected antimalarial medicines circulating in Madagascar, Senegal, and Uganda. *Geneva, Switzerland: WHO.[Google Scholar]*.
40. Chikowe I, Osei-Safo D, Harrison JJ, Konadu DY, Addae-Mensah I, 2015. Post-marketing surveillance of anti-malarial medicines used in Malawi. *Malar J 14:* 127.
41. Khuluza F, Kigera S, Jähnke RW, Heide L, 2016. Use of thin-layer chromatography to detect counterfeit sulfadoxine/pyrimethamine tablets with the wrong active ingredient in Malawi. *Malar J 15:* 215.
42. Khuluza F, Kigera S, Heide L, 2017. Low prevalence of substandard and falsified antimalarial and antibiotic medicines in public and faith-based health facilities of southern Malawi. *The American journal of tropical medicine and hygiene 96:* 1124.
43. Sidibé OI, 2011. Contrôle de qualité des médicaments antipaludiques dans sept régions (O7) administratives du Mali par Chromatographie sur Couche Mince (CCM): Opérationnalisation des kits minlabs.
44. inJos Metropolis CPT, 2001. Chemical Equivalence of Different Brands of Chloroquine Phosphate Tablets inJos Metropolis.
45. Bamiro O, Odeniyi M, Idowu O, Jaiyeoba K, 2004. Physicochemical equivalence of chloroquine phosphate tablets.
46. Aina B, Tayo F, Taylor O, 2007. Quality of chloroquine dosage forms in Lagos State General Hospitals, Nigeria. *J Pharm Pharmacol 59:* 119.
47. Idowu O, Apalara S, Lasisi A, 2006. Assessment of quality of chloroquine tablets sold by drug vendors in Abeokuta, Nigeria. *Tanzania Journal of Health Research 8:* 45-48.
48. Onwujekwe O, Kaur H, Dike N, Shu E, Uzochukwu B, Hanson K, Okoye V, Okonkwo P, 2009. Quality of anti-malarial drugs provided by public and private healthcare providers in south-east Nigeria. *Malar J 8:* 22.
49. Nnamdi JA, Arhewoh IM, Okhamafe AO, Enato EF, 2009. Evaluation of the pharmaceutical quality of some quinine preparations sold in Nigeria. *Med Princ Pract 18:* 193-197.
50. Omeje E, 2008. Evidence for the spectroscopic determination of Artesunate in dosage form. *J Vector Borne Dis*.
51. Esimone C, Okoye FB, Onah B, Nworu C, Omeje E, 2008. In vitro bioequivalence study of nine brands of artesunate tablets marketed in Nigeria.
52. Odunfa O, Adegoke O, Onaga I, 2010. Pharmaceutical equivalence of some commercial samples of Artesunate and Amodiaquine tablets sold in Southwestern Nigeria.
53. Ochekpe N, Agbowuro A, Attah S, 2010. Correlation of price and quality of medicines: assessment of some artemisinin antimalarials in Nigeria based on GPHF minilab. *Int J Drug Dev Res 2:* 211-218.
54. Adegbite A, Adegbolagun O, 2011. Evaluation of the physicochemical equivalence of three brands of commercially available quinine sulphate tablets from South Western part of Nigeria. *Afr Health Sci 11:* 197.

55. Ehianeta T, Williams B, Surakat J, Mohammed N, Anyakora C, 2012. Quality survey of some brands of artesunate-amodiaquine in Lagos drug market. *Afr. J. Pharm. Pharmacol 6:* 636-642.
56. Mathew A, 2015. PHYSIOCHEMICAL PROPERTIES OF SOME PAEDIATRIC FORMULATIONS OF ARTEMETHER-LUMEFANTRINE PRESCRIBED FOR UNCOMPLICATED PLASMODIUM FALCIPARUM MALARIA. *International Journal of Current Research and Review 7:* 41.
57. Izevbekhai O, Adeagbo B, Olagunju A, Bolaji O, 2017. Quality of artemisinin-based antimalarial drugs marketed in Nigeria. *Trans R Soc Trop Med Hyg 111:* 90-96.
58. Adeyemi D, Ogochukwu S, Johnson O, Akinleye M, 2018. Quality assessment of twenty brands of sulfadoxine-pyrimethamine tablets marketed in Mainland. *Journal of Chemical Society of Nigeria 43*.
59. Hassan IA, Adegbola AJ, Soyinka JO, Onyeji CO, Bolaji OO, 2020. Post-marketing surveillance of quality of artemether injection marketed in southwest Nigeria. *The American Journal of Tropical Medicine and Hygiene 103:* 1258.
60. Kayumba P, Risha P, Shewiyo D, Msami A, Masuki G, Ameye D, Vergote G, Ntawukuliryayo J, Remon JP, Vervaet C, 2004. The quality of essential antimicrobial and antimalarial drugs marketed in Rwanda and Tanzania: influence of tropical storage conditions on in vitro dissolution. *J Clin Pharm Ther 29:* 331-338.
61. Habyalimana V, Mbinze Kindenge J, Kalenda Tshilombo N, Dispas A, Yemoa Loconon A, Sacré P-Y, Widart J, De Tullio P, Counerotte S, Kadima Ntokamunda J-L, 2015. Analytical tools and strategic approach to detect poor quality medicines, identify unknown components, and timely alerts for appropriate measures: case study of antimalarial medicines. *American Journal of Analytical Chemistry 6*.
62. Habyalimana V, Mbinze Kindenge J, Yemoa Loconon A, Kadima Ntokamunda J-L, Hubert P, Marini Djang'Eing'A R, 2017. Simple LC isocratic methods development, validation, and application in the analysis of poor quality antimalarial medicines. *American Journal of Analytical Chemistry 8*.
63. Smine A, 2002. *Antimalarial Drug Quality in Senegal*. Available at: http://pdf.usaid.gov/pdf_docs/PNACW987.pdf. Accessed.
64. Jande M, Ngassapa O, Kibwage IO, 2000. Quality of sulfadoxine/pyrimethamine tablets marketed in Dar es Salaam, Tanzania. *The East and Central African Journal of Pharmaceutical Sciences 3:* 20-24.
65. Minzi O, Moshi M, Hipolite D, Massele A, Tomson G, Ericsson Ö, Gustafsson L, 2003. Evaluation of the quality of amodiaquine and sulphadoxine/pyrimethamine tablets sold by private wholesale pharmacies in Dar Es Salaam Tanzania. *J Clin Pharm Ther 28:* 117-122.
66. Risha PG, 2004. Quality of essential drugs on the Tanzanian market: Influence of tropical climate on in vitro dissolution and bioavailability.
67. Risha PG, Msuya Z, Clark M, Johnson K, Ndomondo-Sigonda M, Layloff T, 2008. The use of Minilabs® to improve the testing capacity of regulatory authorities in resource limited settings: Tanzanian experience. *Health Policy 87:* 217-222.
68. Kaur H, Goodman C, Thompson E, Thompson K-A, Masanja I, Kachur SP, Abdulla S, 2008. A nationwide survey of the quality of antimalarials in retail outlets in Tanzania. *PLoS One 3:* e3403.
69. Kaale E, Manyanga V, Chambuso M, Liana J, Rutta E, Embrey M, Layloff T, Johnson K, 2016. The quality of selected essential medicines sold in accredited drug dispensing outlets and pharmacies in Tanzania. *PLoS One 11:* e0165785.
70. Osei-Safo D, Agbonon A, Konadu DY, Harrison JJEK, Edoh M, Gordon A, Gbeassor M, Addae-Mensah I, 2014. Evaluation of the Quality of Artemisinin-Based Antimalarial Medicines Distributed in Ghana and Togo. *Malar Res Treat 2014:* 806416.
71. Ogwal-Okeng J, Okello D, Odyek O, 1998. Quality of oral and parenteral chloroquine in Kampala. *East Afr Med J 75:* 692-694.

72. Ogwal-Okeng JW, Owino E, Obua C, 2003. Chloroquine in the Ugandan market fails quality test: a pharmacovigilance study. *Afr Health Sci 3:* 2-6.
73. Loyce N, 2018. *Survey of Antimalarial medicines and pharmacopoeial quality of Artemether-Lumefantrine tablets sold in private pharmacies and drug shop in Tororo district*. Available at: http://makir.mak.ac.ug/bitstream/handle/10570/6600/NAKALEMBE%20LOYCE%20MSC%20THESIS.pdf?sequence=1&isAllowed=y. Accessed.
74. Ndagije HB, Kiguba R, Manirakiza L, Kirabira E, Sserwanga A, Nabirye L, Mukonzo J, Olsson S, Spinewine A, D'Hoore W, 2020. Healthcare professionals' perspective can guide post-marketing surveillance of artemisinin-based combination therapy in Uganda. *Malar J 19:* 63.
75. Alutuli L, 2012. Evaluation of the Quality of Artemether/Lumefantrine, Sulphadoxine/Pyrimethamine and Quinine Sulphate Tablets in Public and private Health Institutions in Lusaka District.
76. Guo S, He L, Tisch DJ, Kazura J, Mharakurwa S, Mahanta J, Herrera S, Wang B, Cui L, 2016. Pilot testing of dipsticks as point-of-care assays for rapid diagnosis of poor-quality artemisinin drugs in endemic settings. *Trop Med Health 44:* 15.
